# In silico clinical trials of BiTE expression by oncolytic viruses reveal the impact of patient heterogeneity on dosage protocol

**DOI:** 10.64898/2026.07.02.26357107

**Authors:** Adrianne L. Jenner, Robyn P. Araujo, Noa L. Levi, Guy Ungerechts, Christine E. Engeland, Johannes P.W. Heidbuechel

## Abstract

Immunotherapies have become a transformative therapeutic strategy for many cancer types in recent years. Bispecific T-cell engagers (BiTEs) are one promising immunotherapy that enhances cellular antitumour immunity by redirecting T cells towards cancer cells. Recent evidence suggests that BiTE efficacy can be augmented by encoding BiTEs in oncolytic measles virus vectors (MV-BiTE). Infection of cancer cells with MV-BiTE causes the local production of BiTEs and has shown safety and efficacy in murine tumour models. However, whether the observed efficacy of this treatment will translate to a heterogeneous human population is unknown. In this work, we generate an *in silico* clinical trial of MV-BiTE therapy using a system of ordinary differential equations. We capture potential heterogeneity of individual patients using variability in *in vivo* tumour volume and change in baseline (%) measurements from a Phase II clinical trial. In lieu of human MV-BiTE data, we use the oncolytic virus talimogene laherparepvec (T-VEC) as a surrogate oncolytic virus carrying an immunostimulatory payload. Our predictions imply that the main drivers of heterogeneity are the underlying effector T cell killing rate and BiTE pharmacokinetics. Furthermore, we find that if individuals are classified as non-responders to the Phase II T-VEC clinical protocol, they may respond to more frequent administration of lower dosages. This work highlights how *in silico* clinical trials can provide predictions for novel therapeutics to generate hypotheses and guide the design of treatment schedules for clinical translation.

**Author summary:** The immune system has the ability to kill cancer cells; however, cancer cells are able to resist immune cell-mediated killing. A new therapy uses modified measles viruses to activate the immune system against cancer. These viruses are modified with bispecific T-cell engagers (BiTEs) which assist immune cells in targeting and removing cancer cells. While the potential success of this treatment has been demonstrated in mouse models, it is yet to be verified in a human cohort. In this work, we use mathematical and computational simulations to examine how patient-to-patient variability might affect the success of this treatment. We compare our model predictions to data from a clinical trial and find that virtual individuals in the simulation that are classified as non-responders to the standard protocol would likely respond better to more frequent administrations of lower dosages. The work presented here generates hypotheses for how individuals in a human cohort may respond, however, more work is required to verify these results in humans.

## Introduction

Over the past few decades, the treatment of cancer has significantly improved with the introduction of immunotherapies [1]. Broadly speaking, immunotherapies work by eliciting the activity of the patient’s own immune system to recognise and destroy cancer cells. Within the field of immunotherapies, one treatment avenue has been T cell retargeting [2, 3] which aims to bring cytotoxic T lymphocytes into contact with target tumour cells so that cytotoxic responses can be triggered. Bispecific T cell engager^@^ (BiTE) molecules are a recently introduced class of immunotherapeutics that mediate T cell retargeting [2, 4, 5]. Similar to chimeric antigen receptor (CAR) T cell therapy [6], these molecules induce T cell binding to tumour cells by forming cytolytic synapses between T cells and tumour cells, irrespective of T cell receptor specificity and independent of co-stimulation. Unlike standard antibodies, BiTEs have dual antigen specificity, allowing them to bind to two unique antigens and thereby facilitate targeted cell-to-cell interactions [2, 3]. BiTEs are designed to target the T cell antigen CD3 and a tumour-specific antigen simultaneously and promote the cytotoxicity of T cells via crosslinking with tumour cells [4].

Bispecific T cell-recruiting antibodies have been approved for the treatment of haematological malignancies, such as CD19-targeting blinatumomab, approved first-in-class in 2014, while many more are in clinical trials [7–11]. Unfortunately, limited bioavailability and severe toxicities have hampered broader clinical application, especially against solid tumours [12]. Fortunately, oncolytic viruses (OVs) can serve as vectors to deliver BiTEs to solid tumours [12–19], reducing systemic toxicity issues (Fig 1). OVs are naturally occurring or genetically modified viruses which preferentially infect and replicate in tumour cells. Following viral replication, the cell lyses (bursts), releasing the newly generated virions. OVs stimulate an antiviral immune response as well as an antitumour immune response [20]. Potential synergistic effects of BiTE-encoding OVs (OV-BiTEs) result from the inherent ability of the immune system to respond to viral infections, and in doing so, promote the recruitment of T cells to the tumour site which allows for BiTE engagement. Efficacy of the OV-BiTE approach against solid tumours has been postulated by several groups [21–24]. In particular, a recent review [12] highlighted that further demonstration of the applicability and efficacy of OV-BiTEs in a clinical setting is needed as well as investigation of combination therapies [15, 24].

**Fig 1.**
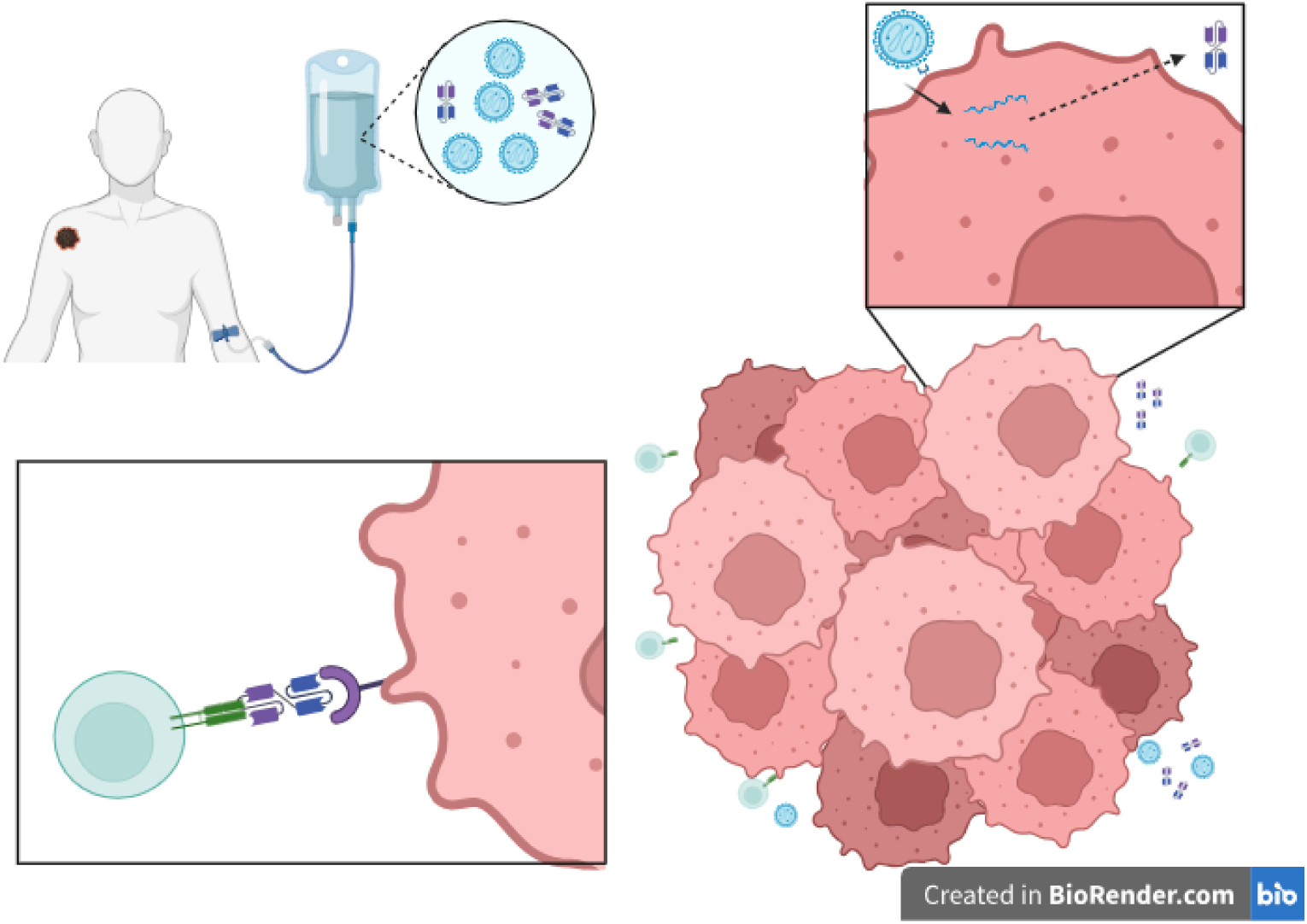
Oncolytic measles viruses encoding bispecific T cell engagers (MV-BiTEs) in the treatment of cancer. MV-BiTEs are administered intratumourally. In pre-clinical studies, due to the virus production method, the suspension may also contain free BiTEs. MV-BiTE-infected tumour cells express and secrete BiTE antibodies. Infected cells ultimately die through tumour cell lysis. T cells are recruited by immune cells sensing the viral infection and by the presence of BiTEs. Simultaneous association of BiTEs with T cells and tumour antigens mediates T cell cytotoxicity against non-infected tumour cells as a bystander effect. Created in BioRender. Engeland, C. (2026).

Mathematical modelling of novel cancer treatments has been a prominent resource in biomedical and cancer research for some time [25, 26]. In particular, a specific focus has been on using mathematics to improve oncolytic virotherapies and immunotherapies [19, 24, 27–32]. Deterministic systems of ordinary differential equations (ODEs) have been employed to investigate novel OV derivatives and the ways in which the resulting antiviral immune response can be harnessed [28, 33–36]. The *in vitro* dose-response relationship of CD3-bispecific antibodies has been examined using a mathematical model [37]. More recently, the first ordinary differential equation system capturing OV-BiTEs was created [27]. While deterministic models are advantageous in their ability to capture mean-field dynamics of a given biological phenomenon, they do not explicitly incorporate heterogeneity that may exist across a cancer patient cohort. To account for this variation in patient characteristics, a virtual clinical trial (VCT) or *in silico* clinical trial can be used [38–40]. There are many examples of VCTs applied to cancer therapies [33, 41–44] and novel techniques are being developed that assist in virtual patient generation and parameter estimation [40, 45, 46].

In this work, we develop an ODE system to represent the action of OV-BiTEs injected intratumourally that subsequently kill tumour cells and stimulate an antitumour immune response. We fit parameters in the model to *in vitro* and *in vivo* measurements for oncolytic measles virus (MV) encoding BiTEs (MV-BiTE) [13]. As there is no human clinical trial data available for MV-BiTE, we generated populations of virtual patients and investigated the efficacy of this therapy in a virtual clinical trial.

We focus our attention on understanding whether an optimal dosage protocol exists for a cohort of heterogeneous individuals, or whether there are individual characteristics that will lead to a more effective treatment. We show that with a simple, cost-effective mathematical simulation we can capture human variations in response to treatment and predict the effectiveness of protocols, which could serve as a useful pre-clinical trial step.

## Materials and methods

### Mathematical model

To better understand MV-BiTE therapy, we generated a system of ordinary differential equations (ODEs) based on previous models for the immune response to immunostimulatory oncolytic viruses [47, 48]. Our model captures a growing uninfected population of tumour cells, *U* (*t*), that become infected by MV-BiTEs, *V* (*t*) and lyse. Inside infected tumour cells, the virus replicates, leading to the production of BiTEs, *B*(*t*), which are released upon cell lysis. The presence of dead cells or cell debris *D*(*t*) stimulates the recruitment of effector CD3^+^ T cells *T* (*t*) to the tumour site. T cells can then induce apoptosis in tumour cells. The presence of BiTEs increases the ability of effector T cells to induce apoptosis in tumour cells. These dynamics are captured in Fig 2, Tables 1 and 2 and the system of ODEs below:

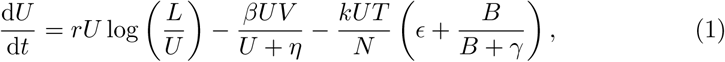

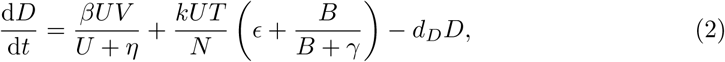

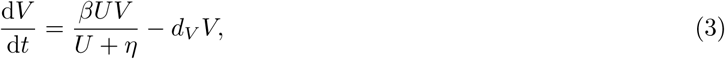

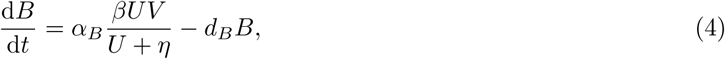

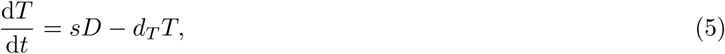

where *t* is time and *N* = *U* + *D* + *T*. To keep the units consistent throughout the model, *V* is in units *cells* which is converted to a quantity of virions by *V* ^∗^ = *α_V_ V*, where *α_V_* has units *virions/cell* and *V* ^∗^ has units virions. For further examples of mathematical models that capture the immune response to oncolytic virotherapy and the resulting antitumour effect, see [28, 34].

**Fig 2.**
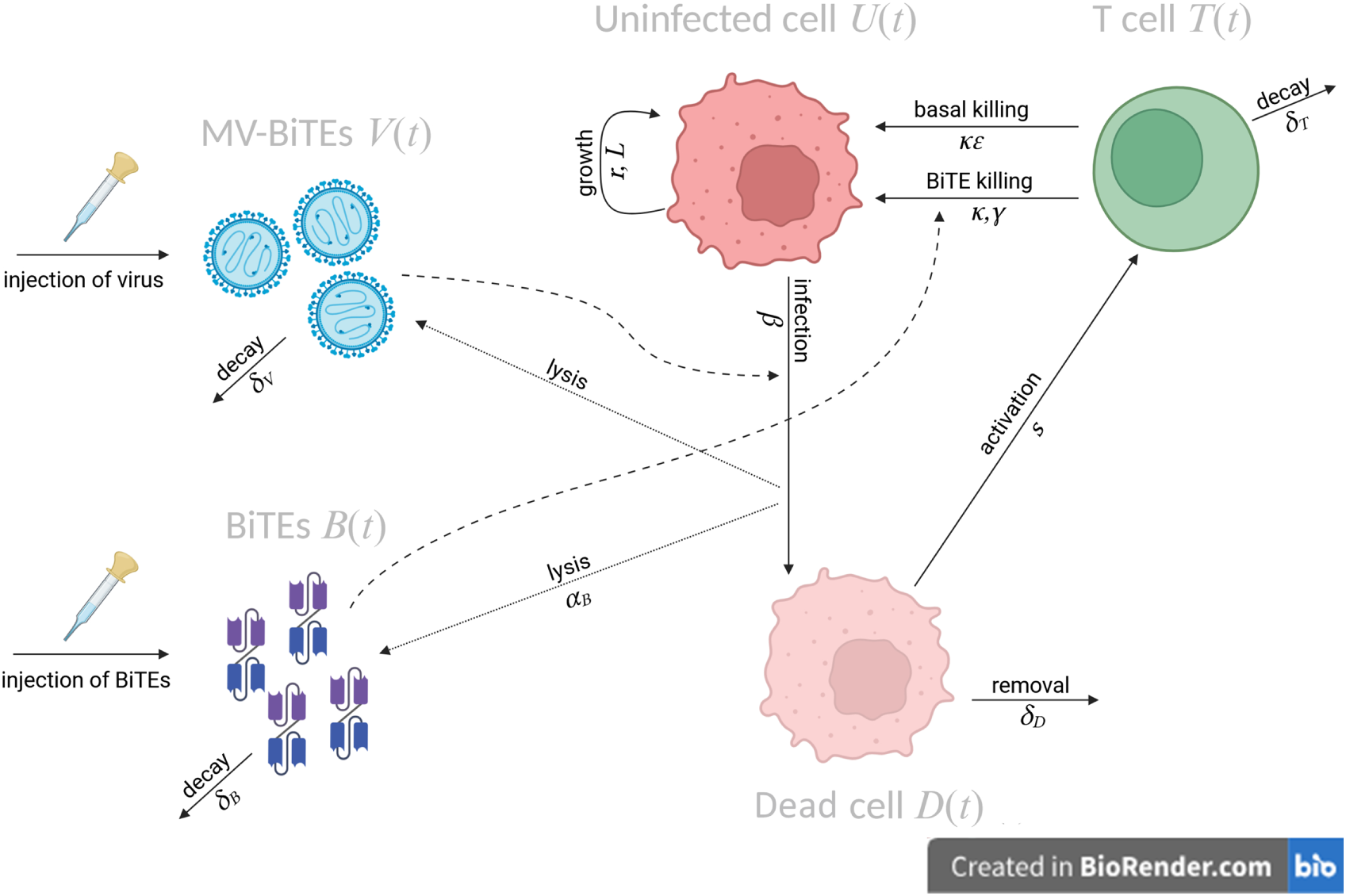
Schematic for the mathematical model in Eqs. 1-5 describing MV-BiTEs. Viruses *V* (*t*), genetically engineered to encode BiTEs, are injected into the system and infect uninfected tumour cells *U* (*t*). These cells eventually die, becoming dead cells, *D*(*t*) and release new viruses and BiTEs *B*(*t*) to the tumour environment. The presence of dead cells activates and recruits CD3^+^ T cells *T* (*t*). T cells induce apoptosis in cancer cells. The presence of BiTEs increases the ability of T cells to induce apoptosis in uninfected tumour cells. Created in BioRender. Engeland, C. (2026)

**Table 1.** Summary of variables in Eqs. 1-5.

| Variable | Units | Description |
| --- | --- | --- |
| $U$ | cells | Not-productively infected (uninfected, eclipse – not secreting BiTEs/virus) pool of cells. |
| $D$ | cells | Productively infected pool of cells producing BiTEs/virus that ultimately die. |
| $V^*$ | virions | Number of oncolytic measles virions, note $V^* = \alpha_V V$. |
| $B$ | $\mu\text{g/ml}$ | BiTE (Bispecific T cell engager) concentration. |
| $T$ | cells | CD3<sup>+</sup> T cells. |

**Table 2.** Parameters for Eqs. 1-5. Estimates were obtained through fitting to *in vitro* and *in vivo* data (Fig 4, S1 Fig,and S2 Fig) with parameters having different values *in vitro* (top row) and *in vivo* (bottom row). Other measurements were estimated based on literature, with their reference given. For the annotation of the experiments below they correspond to the following experiments detailed in the methods: “progeny” - Viral replication kinetics, “viability” - Virus-mediated (onco-)lysis:cell viability, “lysis” – BiTE-mediated (T cell) cytotoxicity: specific lysis, “tumour volume” – *In vivo* tumour volume.

| Parameter | Units | Description | Value | Reference | Experiment |
| --- | --- | --- | --- | --- | --- |
| $r$ | 1/day | proliferation rate of cancer cells | 0.004 | S1 Fig | viability, progeny |
|  |  |  | 0.1321 | Fig 4B | tumour volume |
| $L$ | cells | carrying capacity of tumour | $2.73 \times 10^9$ | Fig 4B | tumour volume |
| $\beta$ | 1/day | infectivity rate | 270.8 | S1 Fig | viability, progeny |
|  |  |  | 25 | S1 Fig | viability, progeny |
| $\eta$ | cells | infection half-effect | $9.04 \times 10^5$ | S1 Fig | viability, progeny |
| $k$ | 1/day | T cell killing rate | 0.1973 | S2 Fig | lysis |
|  |  |  | 0.2567 | Fig 4C-E | tumour volume |
| $\epsilon$ | – | basal T cell killing constant | 0.1716 | S2 Fig | lysis |
|  |  |  | 0.98 | Fig 4C-E | tumour volume |
| $\gamma$ | $\mu\text{g/ml}$ | BiTE half-effect | $4.25 \times 10^{-3}$ | S2 Fig | lysis |
| $d_D$ | 1/day | dead cell disintegration | 5.8 | Fig 4C-E | tumour volume |
| $d_V$ | 1/day | virus decay | 23.11 | S1 Fig | viability, progeny |
| $\alpha_V$ | virions/cell | viruses per cell | 100.56 | S1 Fig | viability, progeny |
| $\alpha_B$ | BiTEs/cell | BiTE production | $4.6 \times 10^4$ | Fig 4C-E | tumour volume |
| $d_B$ | 1/day | decay rate of BiTEs | 2.65 | Fig 4C-E | tumour volume |
| $s$ | 1/day | stimulation of T cells | 28.7 | Fig 4C-E | tumour volume |
| $d_T$ | 1/day | decay rate of T cells | 0.35 | [47] | |

In Eq. 1, tumour cells, *U* (*t*), grow under a Gompertzian tumour growth law [49], under which cells proliferate at rate *r* and the population will grow until a certain carrying capacity *L* is reached. The carrying capacity is a representative of the tumour growth trajectory and does not represent the termination criteria, which may be reached at a smaller value. Tumour cells become infected and undergo lysis at a rate *β*. These infected cells can form syncytia, which we have chosen not to explicitly model here. Infection is modelled as proportional to the concentration of virus and a Michaelis-Menten function with the concentration *η* giving 50% of the maximum infectivity, also known as the half-effect. We introduce this infection term to capture that virus penetration into a tumour is often inhibited spatially either due to the microenvironment or otherwise [50]; see [35, 41, 51] for similar uses of this infectivity formulation. Tumour cells undergo T cell-induced apoptosis at a frequency-dependent rate with constant *k*. For uses of frequency-dependent terms in OV modelling similar to this, see [47, 48, 52]. To model the effect of BiTEs on T cell activity, we assume a baseline T cell apoptosis factor *ɛ*, and that BiTE-mediated killing is an additional factor on top of this baseline killing rate, modelled using a Michaelis-Menten term with half-effect *γ*.

We have removed the intermediate ‘infected’ population that is often present in models of viral infections [28, 34, 47, 53, 54] and instead have direct generation of virus and BiTEs through infection of tumour cells, similar to [55], so that production of BiTEs is modelled continuously over time and not in discrete bursts. We found that introducing the infected cell pool and its specific lysis rate introduced both structural and practical identifiability issues into the model. We also assume infected cells do not replicate after infection, meaning the process of infection is terminal for a cell. In this way, we capture the process of infection, lysis and immune cell recruitment through one population *D*(*t*).

In Eq. 2, we track the accumulation of dead cells or dead cell debris, *D*(*t*), through the induction of lysis by an OV or apoptosis by T cells. To promote the practical identifiability of our model, we chose not to explicitly capture the production of tumour or viral antigens, subsequent activation of antigen-presenting cells, cytokine/chemokine production and then T cell recruitment. As such, we use the accumulation of dead cells and debris as a proxy for this process. Dead cell debris is produced at the same rate that tumour cells die and is removed (decays) from the system at a rate *d_D_*.

In Eq. 3, new viruses are produced through virus infection of tumour cells. These are assumed to decay due to factors such as adaptive immune cell clearance, complement clearance, and sequestration, for example, by the reticuloendothelial system at a rate *d_V_*. As each cell produces *α_V_* virions, we scale the rate of production and decay of virions by *α_V_* to obtain an estimate for the number of virions, *V* ^∗^(*t*).

In Eq. 4, through cell lysis, *α_B_* new BiTEs, *B*(*t*), are released. These BiTEs decay at a rate *d_B_*, i.e., a pharmacokinetic parameter. We assume that these BiTEs consist of both T cell-bound and free BiTEs in the local tumour microenvironment (TME). While BiTEs can be secreted throughout the lifetime of the cell, we found separating the number of BiTEs released at lysis *α_B_*from the amount secreted up to lysis was not structurally identifiable with the given experimental measurements. As such, we assume that any BiTEs released up to lysis are included in the amount secreted at lysis *α_B_*.

In Eq. 5, T cells, *T* (*t*), accumulate at the tumour site based on the presence of dead cell debris at a rate *s*. These cells also decay at a rate *d_T_*. We assume T cells are recruited at a rate *sD*, which can be thought of as a proxy for antigen-presenting cell activation by viral antigens or tumour antigens, downstream stimulation of T cells in the lymph nodes, and tumour-directed T cell chemotaxis.

The model was simulated in MATLAB and solved numerically using *ode45*. Initial conditions are given below:

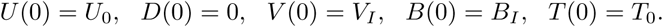

The model was solved piecewise to simulate *n* injections of BiTEs *B*_0_ and virus *V*_0_ at time *t_I_*:

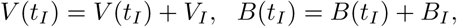

for *I* = 1*, …, n*.

### Structural and practical identifiability

Identifiability is an important component of mathematical modelling as it determines the capacity of a model to extract parameter estimates from data [56–59]. This is particularly important in biology, where parameterised models are used to make predictions about living systems [60, 61]. We refined the model presented in Eqs 1-5 numerous times to minimise identifiability issues. The model was found to be structurally identifiable using the open-source software DAISY [62, 63]. Structural identifiability implies practical identifiability only for infinite data [56]. For finite data samples, there are numerous ways to evaluate practical identifiability [64–66]; for our dataset and model, we have analysed practical identifiability with a global and local sensitivity analysis.

### Details of experimental and clinical measurements

To estimate parameters in Eqs 1-5, we use a range of *in vitro* and *in vivo* experiments from oncolytic measles virus (MV) encoding and not encoding BiTEs [13], as well as a Phase II clinical trial for talimogene laherparepvec (T-VEC) [67]. Vaccine-derived, live attenuated MV vectors were designed based on established reverse genetics systems and were generated to encode BiTEs targeting murine CD3 and human CD20 [68]. For *in vitro* studies and the syngeneic subcutaneous tumour model in C57Bl/6J mice, murine melanoma-derived B16-CD20-CD46 cells stably expressing the BiTE target antigen human CD20 and human CD46 as entry receptor for oncolytic measles virus were used.

#### Viral replication kinetics

Replication kinetics of MV were captured by determining the viral progeny produced in B16-CD20-CD46. 10^5^ cells on 12-well plates were inoculated with the same dose (multiplicity of infection, MOI of 1) of cell infectious units (ciu) of MV encoding mCD3xCD20-BiTE (with one scFv targeting murine CD3 expressed on mouse T cells and the other recognizing the human CD20 antigen) for 12 hours. Inoculum was then replaced by medium. At designated time points, supernatants were collected, samples were frozen and viral progeny was assessed via serial dilution titration assay.

#### Virus-mediated (onco-)lysis: cell viability

Cytotoxic effects of MV were determined in B16-CD20-CD46 cells. The cells were infected with unmodified MV at an MOI of 1 as for the replication assay and cell viability was determined by metabolic XTT assay every 12 hours for 96 hours. Viability was calculated relative to mock-infected cells.

#### BiTE-mediated (T cell) cytotoxicity: specific lysis

T cell cytotoxicity mediated by BiTEs was determined in B16-CD20-CD46 tumour cells expressing the BiTE target antigen co-cultured with primary, unstimulated immune effector cells obtained from mouse spleens at varying BiTE concentrations. Three experiments with independently produced mCD3xCD20 BiTE batches purified from the supernatant of MV-mCD3xCD20-infected (Vero producer) cells were performed. LDH release was determined at 44h after setting up the co-cultures and specific tumour cell lysis was calculated relative to chemically lysed maximum release controls following subtraction of background values taken from target cell only and effector cell only controls, as described in Heidbuechel and Engeland [12] and Speck *et al.* [13].

### *In vivo* tumour volume

For *in vivo* experiments, C57/BL6J mice were injected with 10^6^ B16-CD20-CD46 cells s.c. After tumour engraftment, mice were stratified to respective treatment groups according to tumour size and treated with intratumoural injections. The experimenter was blinded to treatment groups. Treatment conditions included carrier fluid only (mock), purified BiTE (mCD3xCD20, as in the LDH release assay), unmodified measles virus (MV), and MV encoding CD20-targeting BiTE (MV-mCD3xCD20). The injection protocol was 5 intratumoural injections administered on consecutive days of 100*µ*L volume each containing 10^6^ cell infectious units (i.e., infectious virus as assessed by titration). In the respective “BiTE only” treatment group, 2.17*µ*g of purified BiTE (in 100*µ*L suspension) was injected 5x instead of virus, which corresponds to what was found to be present in 100*µ*L of MV-BiTE suspension via flow cytometry in a quantitative binding assay. Note that the estimate of 2.17*µ*g would depend on the injection and tumour volume, however, for simplicity we have assumed this as a fixed amount throughout our simulations.

Caliper measurements were performed every three days to determine tumour volume. Tumour volumes were calculated as [larger diameter *×* smaller diameter *×* smaller diameter]/2. All experiments were conducted with non-immunocompromised wild-type C57BL/6 mice with endogenous CD3^+^ T cells. Intratumoural T cell abundance is known to be quite low in the B16 model [69] and increased upon treatment (more so in the MV-BiTE vs MV experiment as assessed by flow cytometry). If ulceration presented in the mice as the tumours formed, these mice were not included in this study.

#### Phase II Clinical Trial measurements for change from baseline (%)

To evaluate the function of the model in a clinical setting, data from a Phase II T-VEC trial [67] was used. T-VEC is a herpes simplex virus type 1-derived oncolytic immunotherapy designed to selectively replicate within tumours and encode granulocyte-macrophage colony-stimulating factor (GM-CSF) to enhance systemic antitumour immune responses. Eligible patients were age *≥* 18 years with unresectable stage IIIB-IV melanoma. T-VEC was administered by intralesional injection following the dosing regimen described in the registrational Phase III OPTiM study [70], which was a first dose of T-VEC of 10^6^ pfu/ml, followed by T-VEC doses of 10^8^ pfu/ml 3 weeks after the first dose and then once every 2 weeks. Treatment continued for *≥* 6 months from initial dosing provided treatment was tolerated.

### Parameter estimation

To estimate parameters in the model, a series of hierarchical parameter fits were conducted to *in vitro* and *in vivo* measurements using MATLAB’s inbuilt non-linear least squares optimisation algorithm *lsqnonlin* and the inbuilt multistart algorithm *multistart*. The viral infection and replication parameters *β, η, d_V_*and *α* were estimated using the *in vitro* viral progeny and cell viability experiments, see in S1 Text and S1 Fig. The BiTE and T cell killing parameters *ɛ, k*, and *γ* were obtained through fitting the specific lysis (%) data, S2 Fig. The *in vivo* tumour growth rate without treatment, *r* and *L*, were obtained through fitting to the control data set, assuming that 10^6^ cells were injected at day 0 and 1mm^3^ = 10^6^ cells [71].

Given the initial parameter estimates from the *in vitro* experiments, we then recalibrated parameters to the *in vivo* measurements. We simultaneously fit the model to the purified BiTEs, MV and MV-BiTEs measurements to obtain *β, k, ɛ, d_D_, α_B_, d_B_*, and *s*. The decay rate of T cells was taken from data fitting, *d_T_*= 0.35/day, [48]. The time from initiation of cell apoptosis to completion can occur as quickly as 2-3 hours. If we assume that debris can remain for another 2 hours after this, this gives a dead cell clearance rate of *d_D_*= 4.8/day [72]. For fitting the *in vivo* data: *B*(0) = 2.17*µ*g, and *V* (0) = 10^6^ cells. Further details of each fit are provided in the Supplementary Material.

### Sensitivity analysis

We evaluated the sensitivity of our model predictions to perturbations to the obtained parameter estimates through a local and global sensitivity analysis. We conducted a pairwise local sensitivity analysis around the fitted parameter values, see Supplementary Material and S3 Fig. We also conducted a global sensitivity analysis by running a Latin Hypercube Sampling and calculating the Pearson’s Correlation Coefficient (PCC) for each parameter with the final tumour size. Other sensitivity analysis methods, such as eFAST, MeFAST or Sobol, would also have been appropriate here [73].

### *In silico* clinical trial

Virtual clinical trials (VCT), also known as virtual patient cohorts, phase i trials, digital twins, and *in silico* clinical trials attempt to integrate the heterogeneity of actual patient responses and preclinical studies through a cohort of virtual patients or individuals [33, 38–44, 74].

#### Virtual patient generation from *in vivo* data

There are many ways to generate virtual patients [41, 45, 46] and in this work, we created virtual cohorts using two different methods employed by previous works [41, 44, 45]. We first created a cohort of patients that matched individual *in vivo* mouse tumour growth trajectories using acceptance-rejection sampling (Fig 3: Method (1)). We sampled 250,000,000 parameter sets *θ* = *k, ɛ, d_B_, d_T_, s, η, β* from uniform distributions centered around the fitted parameter *p*^ values with bounds [0.001, 5]. We took the 400 parameter sets that gave the minimum residual to the individual tumour volume measurements, labelled “m1” to “m8”. The second method we used to generate a virtual cohort, was a similar approach to that developed by Kim *et al.* [44]. We converted the change from baseline measurements for the clinical trial data to a probability density function (PDF) for the final tumour size *x* (Fig 3: Method (2)). We then sampled 400 patients from this PDF and optimised the set of parameters to obtain an estimate for the final model size that matched the sampled patient using a genetic algorithm (MATLAB’s *ga*). This gave us a distribution of final tumour sizes and corresponding parameter values that matched the PDF obtained from the clinical data.

**Fig 3.**
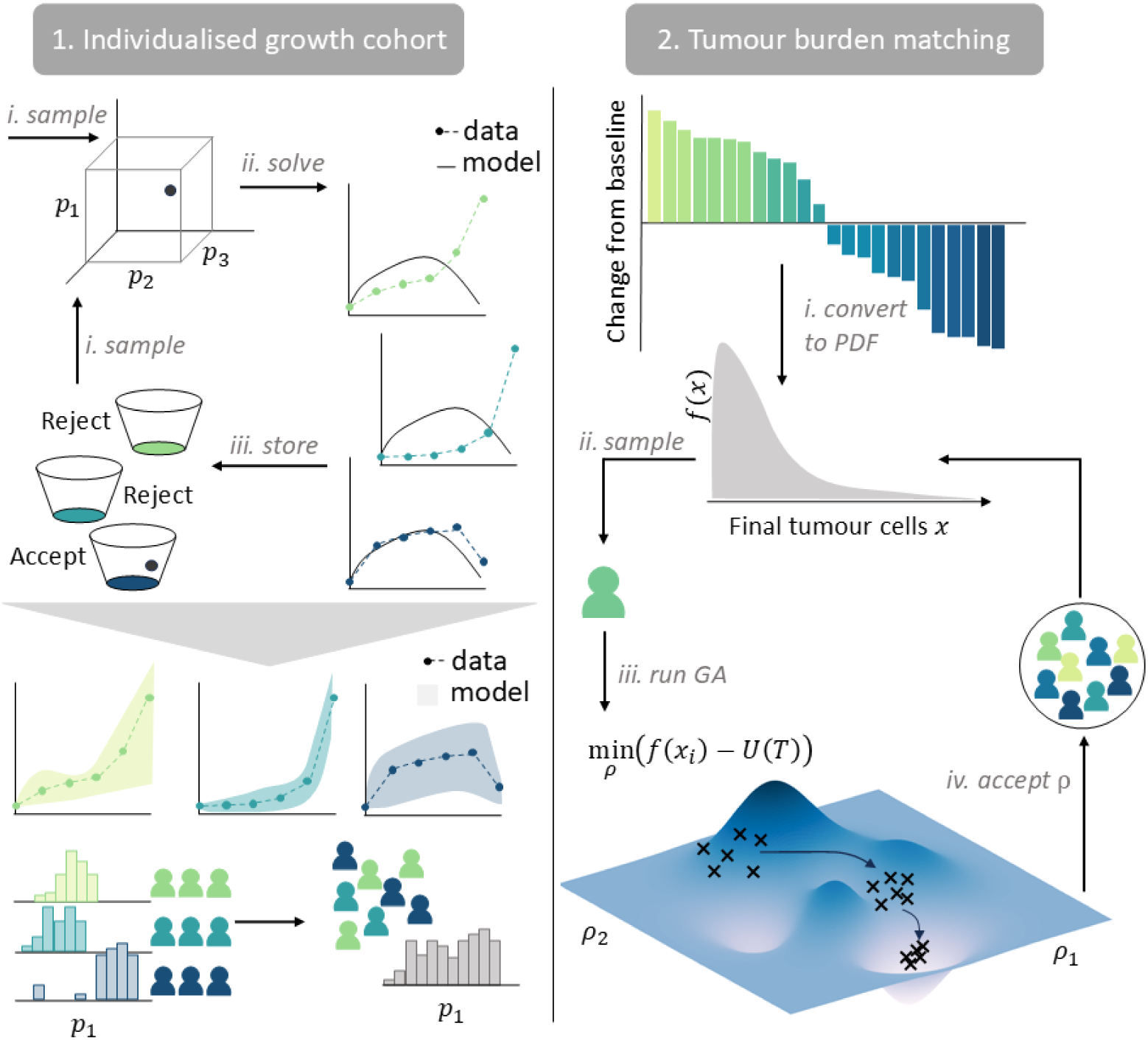
Illustration of the two virtual clinical trial generation methods: Method (1) individualised growth cohort and Method (2) tumour burden matching. Method (1) investigates heterogeneity arising in the *in vivo* measurements by matching the model to individual mouse tumour growth trajectories. Parameters are sampled from uniform multi-dimensional parameter distributions, and an acceptance-rejection algorithm is implemented, accepting samples that align closest with the data. Once a cohort of *N* samples for each individual tumour growth trajectory have been sampled, the distribution for these parameters, e.g. *p*_1_, can be compared across the different trajectories. Method (2) generates the virtual cohort through converting measurements from a clinical trial measuring the change from baseline (CFB) to a probability density function (PDF) *f* (*x*) for the final tumour size *x*. From this PDF, *N* individual final tumour sizes are sampled and a genetic algorithm is used to determine the optimal parameter combination for the model to match the CFB (%) data.

#### Virtual patient generation from Phase II clinical trial

In the Phase II T-VEC clinical trial, patients were required to have melanoma lesions appropriate for intralesional injection, defined as *≥* 1 lesion with a diameter of *≥* 10 mm or multiple lesions with a combined diameter of *≥* 10mm. Previous work by Dessinioti *et al.* [75] reported a median melanoma diameter of 13mm, with quartiles 9 - 20mm, and 10.6% of tumours below 6mm. Using these statistical summaries, we parameterised a gamma distribution and obtained a shape and scale parameter of 3.09 and 4.91 respectively (see Supplementary Information). To convert from tumour diameter to cell count, we assumed a spherical tumour volume, which gives 1, 150.35mm^3^. We then assumed that 1mm^3^ = 10^6^ cells which hl, for an initial tumour diameter of 13mm, gives an initial cell count of *U* (0) = 1.15 *×* 10^9^ cells. For the first virtual cohort investigation of the clinical trial measurements, we sampled 400 tumour diameters from the parameterised gamma distribution and converted them to cell counts which were used as initial conditions for the model. Following this, we fixed the initial cell count to the median tumour diameter, i.e. 13mm, and obtain virtual patients using Method (2) described above.

At baseline, the mean intratumoural CD8+ T cell density was 460 cells/mm^2^ (range 5-3,963) for patients in the clinical trial [67]. If we assume the measurements were taken on 20*µ*m samples, then this would give the approximate density of T cells in a 1mm^3^ sample to be 4.6 *×* 10^4^ cells*/*mm^3^. As both *L* and *η* have cells as units and were estimated *in vivo* these were rescaled to a human context using the new initial number of tumour cells, see the Supplementary Information for explicit calculations. We have chosen to evaluate all patients at an equivalent final time to *T* = 23 weeks, as all treatments were continued for at least 6 months (unless they were not tolerated by the patient) [67]. To confirm the distribution for the change from baseline of the Phase II clinical trial cohort matched the virtual cohort, we used a Kolmogorov-Smirnov test for similarity using Matlab’s function *kstest2*.

## Results

### Parameterisation and sensitivity of a mathematical representation for MV-BiTEs

To calibrate the model, we fit parameters from the system of ODEs in Eqs 1-5 to *in vitro* measurements for viral progeny and cell viability (S1 Fig), and specific lysis (S2 Fig). We assumed the viral infectivity and CD3^+^ T cell killing rate may vary *in vivo* and refit these using the *in vivo* tumour volume experiments (Fig 4A-E). Estimates for parameters can be found in Table 2 with more details in the Materials and Methods and Supplementary Information S1 Text. Qualitatively, we observed good agreement between the data and the model, supported by goodness of fit statistics, Table 3. To investigate the sensitivity of the parameterised model, we conducted a local and global parameter sensitivity analysis (S3 Fig and Fig 4F). The global sensitivity analysis suggested the immune-related parameters *k, ɛ*, and *s* had a negative correlation with the tumour size.

**Fig 4.**
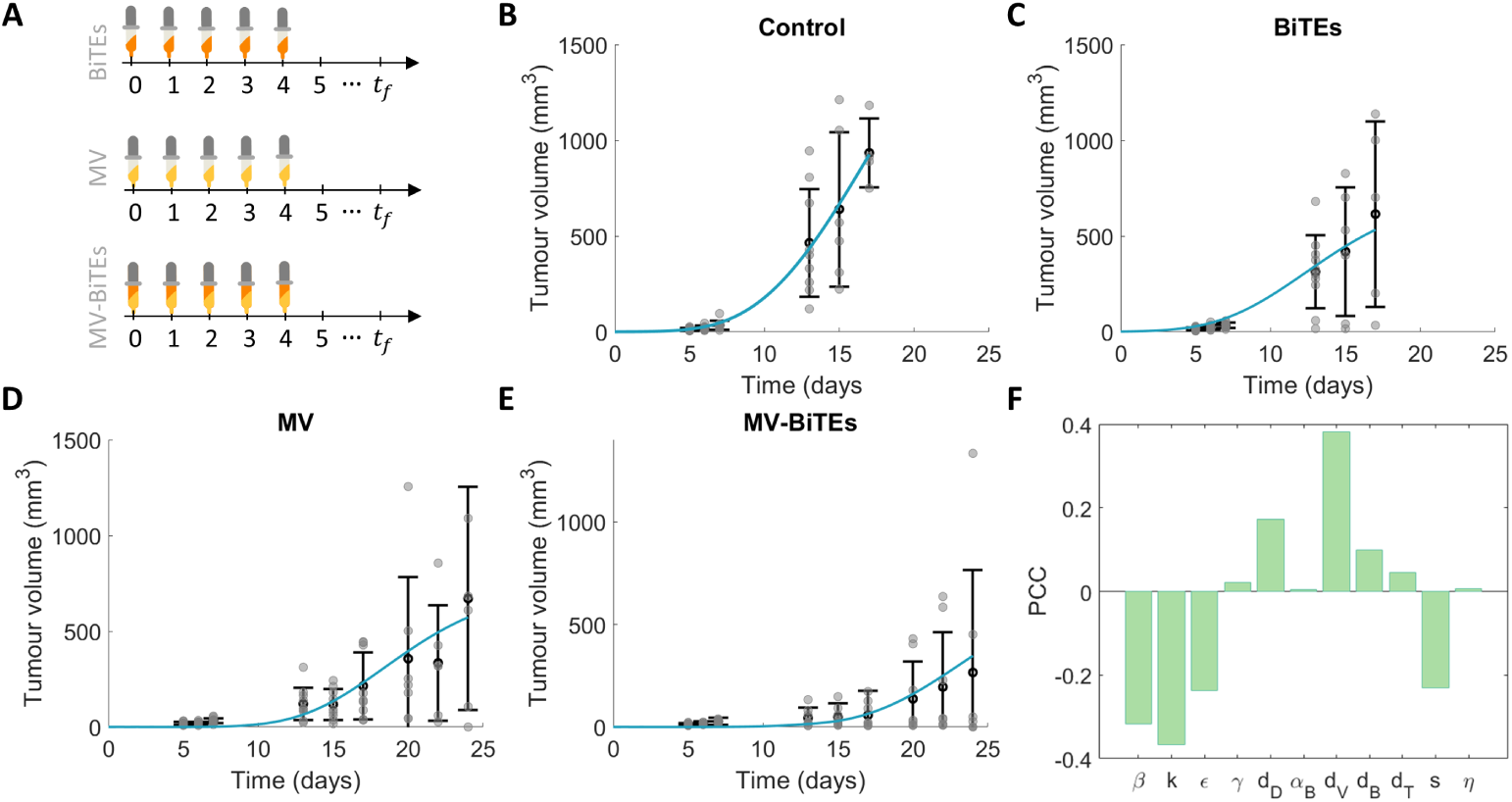
Parameterisation of the model for MV-BiTEs using *in vitro* and *in vivo* experiments. Generation of data is described in the Methods. (A) Experimental protocol for *in vivo* experiments with BiTEs, MV and MV-BiTEs. (B)-(E) Tumour volume *in vivo* in (B) control (mock) and (C) BiTEs only, (D) MV only and (E) MV-BiTEs treatment groups. Black circles with error bars are representative of the mean and standard deviation of the data and grey circles are individual data points. Solid blue lines represent the model’s approximation to the data. (F) Pearson Correlation Coefficient (PCC) for a Latin Hypercube Sampling on [0.01, 2] capturing the sensitivity of the estimated parameters.

**Table 3.** Goodness of fit statistics for model fit to the data in Fig 4, with parameter values in Table 2.

| Experiment | Figure | R-square | Residual |
| --- | --- | --- | --- |
| *in vivo* control | Fig 4D | 0.9988 | $1.84 \times 10^3$ |
| *in vivo* BiTEs | Fig 4E | 0.9892 | $1.06 \times 10^4$ |
| *in vivo* MV | Fig 4F | 0.9466 | $4.22 \times 10^4$ |
| *in vivo* MV-BiTE | Fig 4G | 0.9968 | $1.27 \times 10^4$ |

### Capturing inherent *in vivo* variability in tumour responses

In the *in vivo* experiment, mice exhibited heterogeneous responses to MV-BiTE treatment, with some mice experiencing tumour progression and others experiencing regression (Fig 5). To examine the model’s ability to capture this heterogeneity, we sampled a virtual cohort using Method (1) (Fig 3) for each individual mouse, m1-m8. We obtained 400 parameter sets for each m1-m8 trajectory that gave the smallest residual to the data (Fig 5A-I and S4 Fig). The model was able to capture the heterogeneous range of tumour growths exhibited by the mice upon MV-BiTE treatment.

**Fig 5.**
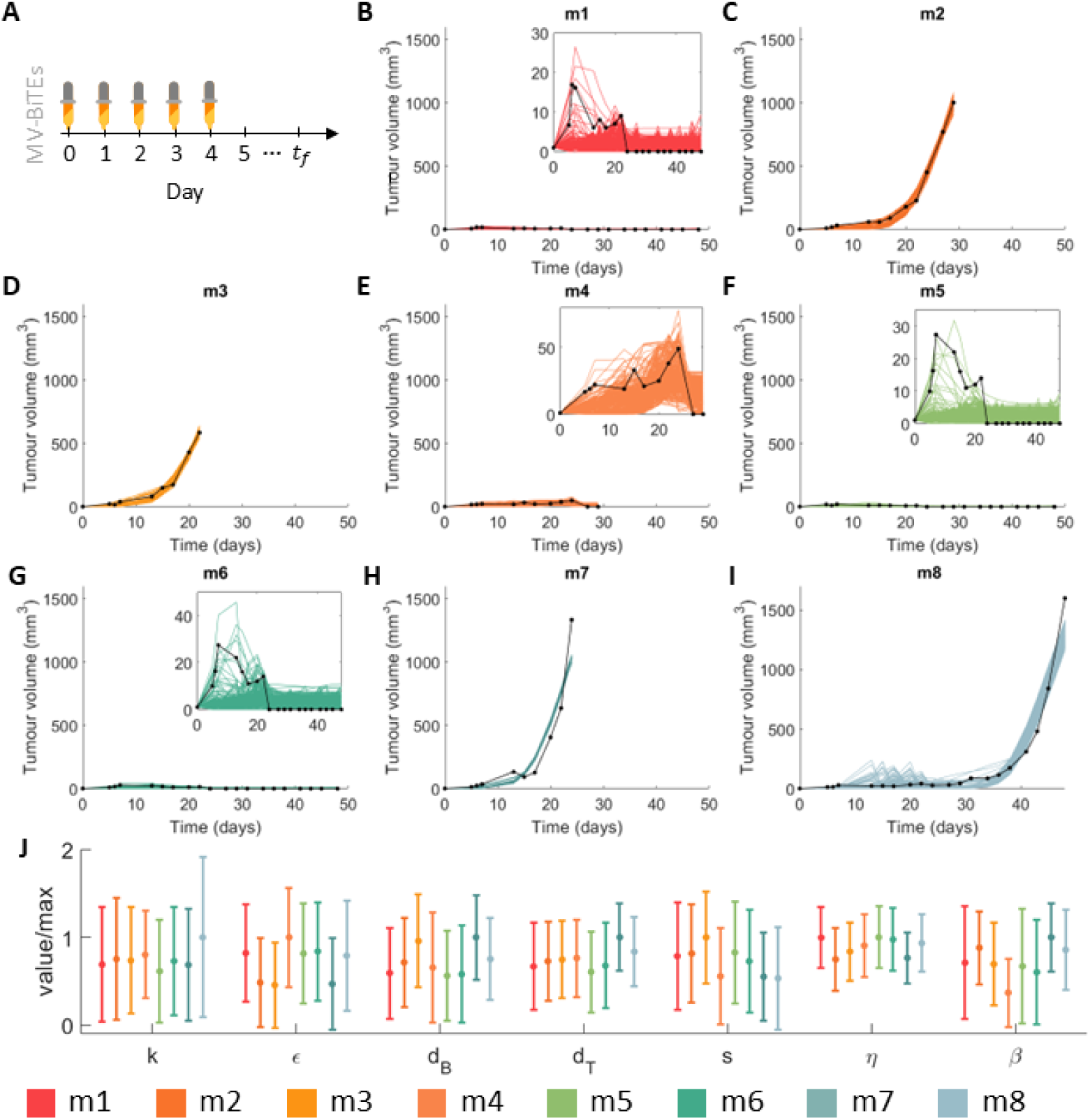
Obtaining model trajectories that match the tumour volume experimental measurements under the treatment schedule in (A). In the experiments, the tumour volume of 8 mice was tracked after treatment with MV-BiTEs with m1 to m8 in (B)-(I). The measurements for the mice are plotted on the same axis and represented by black dots, while the trajectories of the 400 parameter values sampled that gave the lowest residual error are plotted as coloured lines. Zoomed in inserts are provided when the trajectories are hard to visualise on the fixed axis. (J) The mean and standard deviation of the relative value for the parameters for each m1 to m8 are given as an error bar plot. These parameter ranges have been scaled relative to the maximum so they can be seen on the same figure. The corresponding parameter values and pair-wise correlations can be seen in the S4 Fig-S6 Fig.

Variability between the tumour growths is captured by the relative mean and standard deviation for each parameter (Fig 5J and S5 Fig). Noticeable differences in the mean for *β* exist, suggesting that virus administration is largely driving the heterogeneous responses in the mice. Furthermore, changes in *ɛ* and *d_B_*seem to correlate with either tumour growth (m2, m3, and m7) or tumour decline (m1, m4-m6). This suggests that tumour growth or decline could be linked to the efficacy of CD3^+^ T cells and the pharmacokinetics of BiTEs.

Examining correlations between parameters, we found that all individual growths, except m2, had a positive correlation between viral characteristics, *η* and *β* (S6 Fig), i.e., increasing one of these parameters resulted in an increase of the other.

Furthermore, m2 - m6 all had a negative correlation between *k* and *ɛ*, i.e., increases in one parameter observed a decrease in the other. This suggests a link between viral parameters *η* and *β*, immune parameters *k* and *ɛ*, and the response to MV-BiTEs.

### Clustering individual responses into growth, delayed growth and remission

The responses to MV-BiTEs treatment can be grouped into three cohorts: 1. regression (m1, m4, m5, m6), 2. growth (m2, m3, m7,) and 3. delayed growth (m8). We generated data for these three virtual cohorts by sampling 400 parameter sets equally split amongst the mice belonging to that cohort (Fig 6A-C). Clear separation between the response cohorts can be seen in the final tumour size on day 48 (the final measurement across m1-m8) (Fig 6D). Not all trajectories in the growth cohort (cohort 2) are expected to surpass the delayed growth cohort, with some exhibiting small tumour sizes at day 48. This suggests that while the delayed growth cohort may at first seem to fare better compared to the growth cohort, some favourable trajectories are expected in the growth cohort.

**Fig 6.**
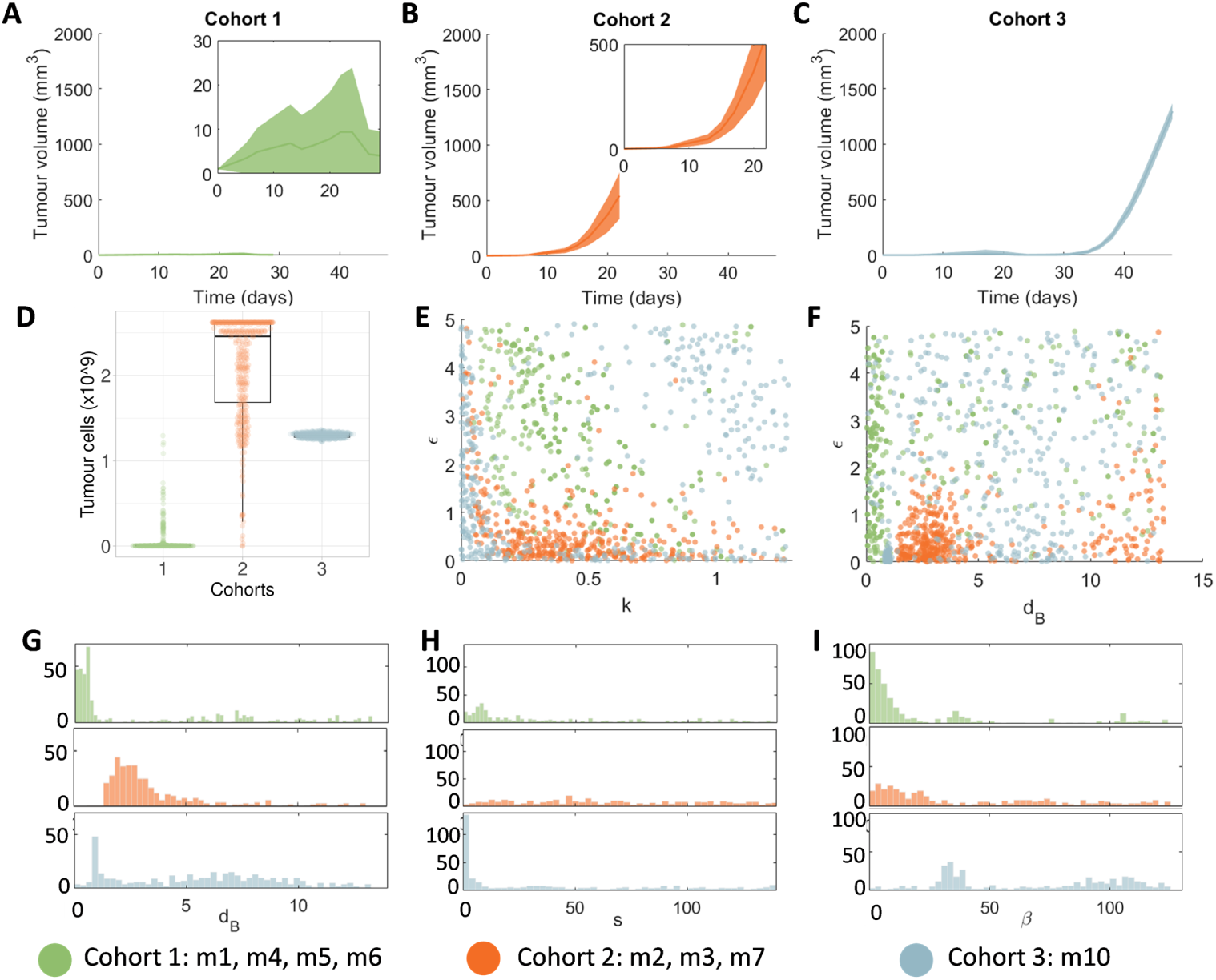
Analysing cohort characteristics. From the mice in Fig 5, three sub-cohorts were generated based on the dynamics in the data: regression (cohort 1), growth (cohort 2), delayed growth (cohort 3). (A-C) Mean tumour volumes and standard deviations for the three cohort values. (D) Final tumour cell number (as calculated for day 48) for each cohort. (E-F) For the three cohorts, the patterns within the parameter values are presented using scatter plots for *ɛ* vs *k* and *d_B_*vs *ɛ*. Points are coloured based on the cohort. (G-I) Histograms for the parameters *d_B_, s*, and *β*. higher *d_B_*, suggesting again that BiTE clearance is a key determinant of the response to MV-BiTEs.

Examining correlations between model parameters across the cohort we see distinctions in the clustering patterns for *k* and *ɛ* (Fig 6E). This suggests that the relationship between the killing rate and the basal killing efficacy are driving the response to MV-BiTE: low basal killing efficacy (*ɛ*) drives the growth cohort (cohort 2) and higher ratios of basal killing efficacy, and killing rates, drive the recovery cohort (cohort 1). Interestingly, there seems to be a bimodal relationship between these two parameters for the delayed growth cohort (cohort 3) suggesting that there are two possible explanations for tumour growth in this cohort. Similarly, there is clustering in the relationship between *d_B_*and *ɛ* (Fig 6F), whereby for small basal killing efficacy and values of BiTE clearance, *d_B_*, between 2 and 7 gives rise to the growth cohort (cohort 2). Comparing single parameter distributions (Fig 6G-I) the regression cohort (cohort 1) has very low *d_B_*and *β* and the growth cohort (cohort 2) has very low *β* but slightly

### Efficacy of administration protocol on variable tumour growths

For the three response cohorts (regression, growth, and delayed growth), we next investigated the effect of varying the MV-BiTEs administration protocol. We varied the number of dosages, *n*, and days between dosages, *τ* (Fig 7A) with the total virus dose kept constant. Variations to the MV-BiTEs protocol had a negligible effect on the regression cohort (cohort 1) (Fig 7B (top row) and S7 Fig), suggesting this cohort will always respond to MV-BiTEs irrespective of the administration protocol. A larger dependence on the protocol is seen for the growth cohort (cohort 2) and delayed growth cohort (cohort 3). The optimal protocol for these cohorts is more injections given further apart.

**Fig 7.**
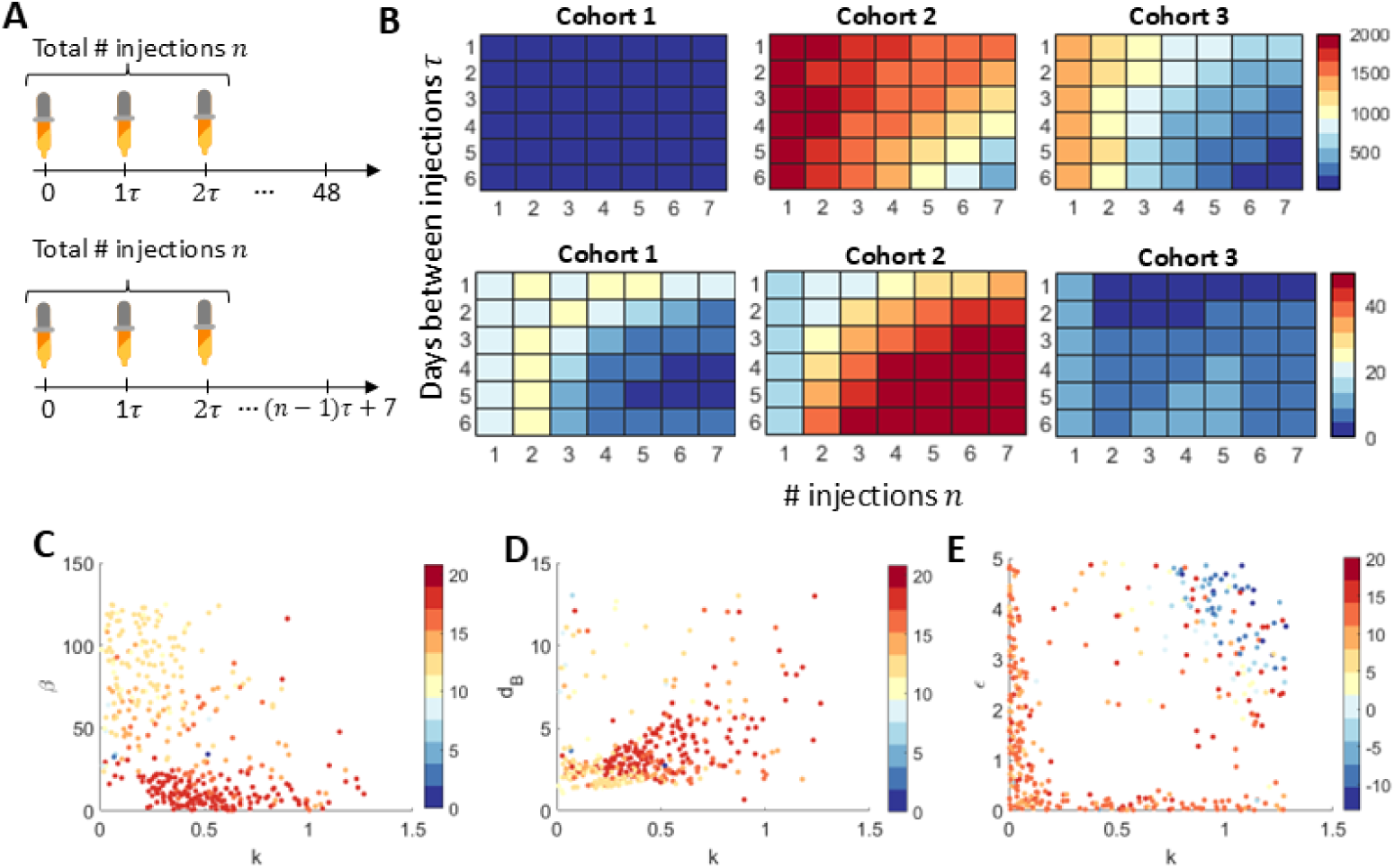
The impact of the treatment protocol on the three cohort responses: a) regression, 2) growth, and 3) delayed growth. (A) Treatment protocols considered varied the number of injections n and days between injections *τ*. (B) The effect of the protocol on the mean tumour size (in mm3) for each cohort at either 48 days (top row) or 7 days after the final injection (bottom row). (C-D) Parameter correlations for cohort 2: (C) *k* vs *β* and (D) *k* vs *d_B_*. (E) Parameter correlations for cohort 3 for *k* vs *ɛ*. All scatter plots are based on 7 injections given 6 days apart with parameter pairs represented as points coloured by the log tumour size at 48 days.

Taking the optimal protocol to be 7 injections 6 days apart, we found that there is clear separation between the best and worst responses in the growth (cohort 2) and delayed growth (cohort 3) cohorts. For instance, in cohort 2, there is a clear separation in the pair-wise relationships between *d_B_* and *β*, and *d_B_* and *k*, respectively (Fig 7C-D). This suggests that we can determine individuals who will respond best to this treatment protocol based on their BiTE pharmacokinetics, viral infectivity and CD3 killing rates. Similarly, in the delayed growth cohort (cohort 3) (Fig 7E) individuals who respond best have high CD3 killing and basal kill rates. We then examined whether this optimal protocol holds under the condition that after treatment ends, we resect the tumour 7 days later (Fig 8B (bottom row)). In this scenario, all protocols are similar as 7 days after treatment, the model predicts all cohorts are responding to MV-BiTEs. The rationale behind this metric is that it helps to evaluate the efficacy of treatment in controlling tumour growth by examining the impact once the treatment is halted and the tumour has time to regrow.

**Fig 8.**
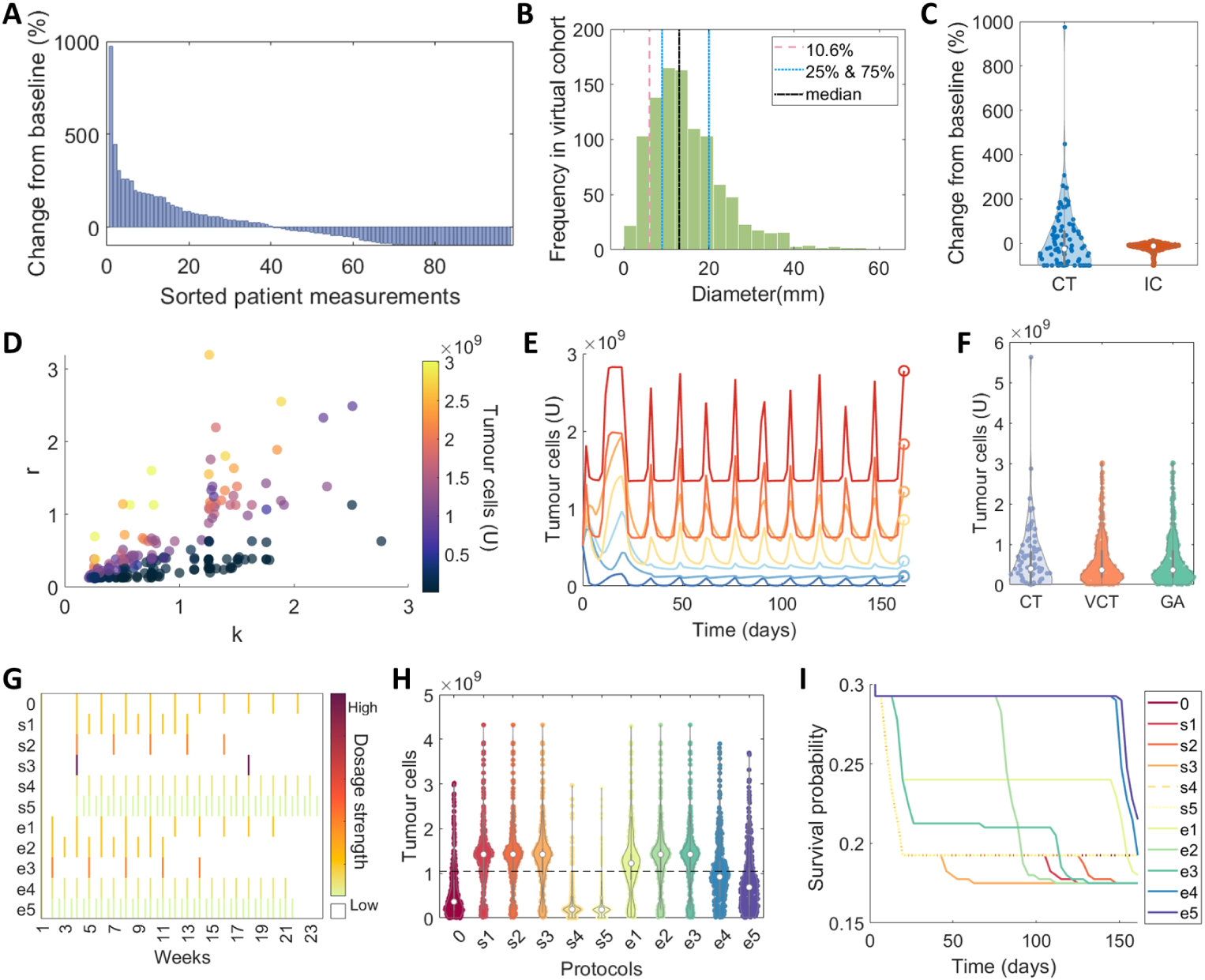
Expanding a virtual patient cohort informed by the Phase II T-VEC clinical trial [67]. (A) Change from baseline (%) for the Phase II T-VEC clinical trial digitized using WebPlotDigitiser [76]. (B) Distribution of initial tumour diameters for 400 virtual patients sampled from a gamma distribution estimated from the summary statistics of Dessinioti *et al.* [75] overlaid as vertical lines: 10.6% (dashed orange), 25% and 75% (blue dotted), and median (black dot dash). (C) Violin plots comparing the change from baseline for the clinical trial (CT) measurements and model simulations from the sampled initial tumour diameters (IC). (D) Scatter plot for parameters *r* vs *k* obtained from optimising the model output to the CT measurements using a genetic algorithm, i.e. Method (2). Each disk represents a patient with the colour representing the tumour cell count. (E) Plot showing the agreement between the model (solid lines) and patient samples (open circles) for 7 virtual patients (colour matched). (F) The distribution for the total number of tumour cells predicted by the model at T=23 weeks and those in the CT, the samples for the 400 virtual patients (VCT), and model predictions using a genetic algorithm (GA).(G) Summary of the original clinical trial dosage protocol and the 10 hypothetical protocols considered (Table 4). Coloured bars represent the strength of a dosage given in a particular week. (H) The final tumour cell counts for varying treatment protocols examined (s1-s5, e1-e5) compared to the original protocol 0. (I) Kaplan-Meier survival curves corresponding to (E) for a threshold of a 120% CFB. Note the survival probability range is plotted on [0.15 to 0.3].

**Table 4.**
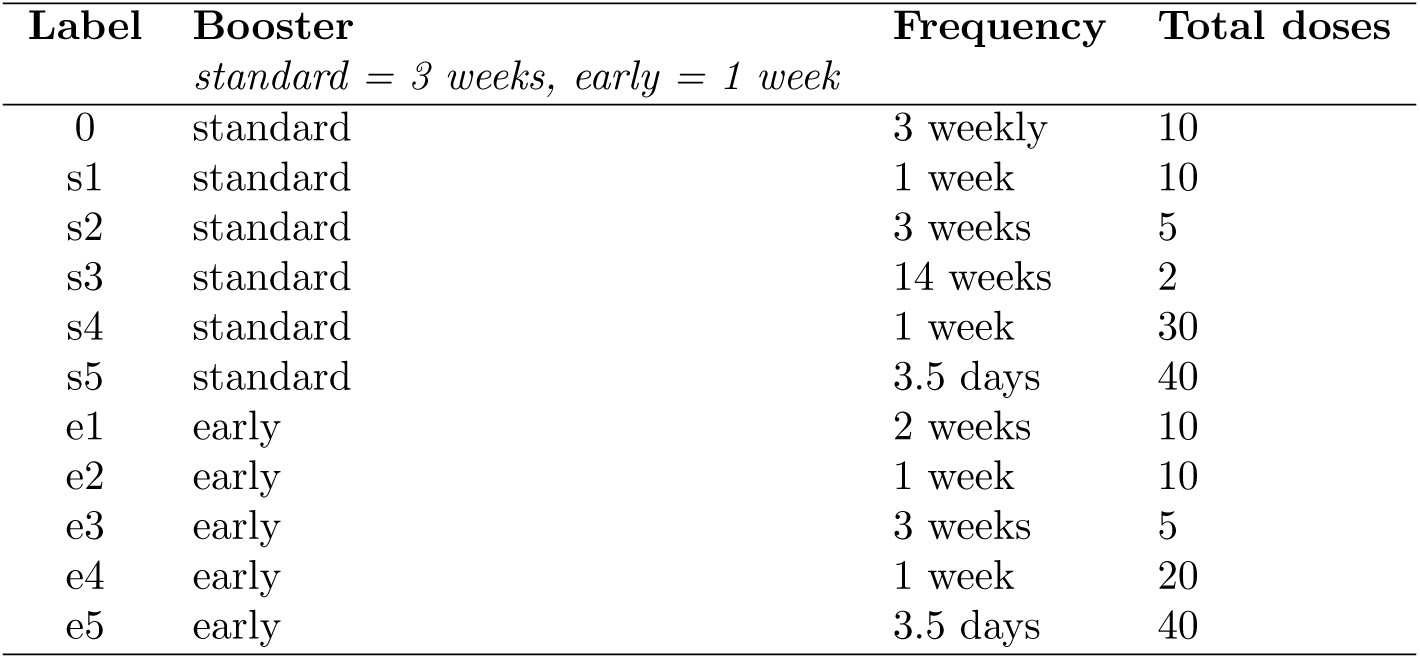
Summary of the standard clinical trial protocol and the 10 hypothetical protocols examined in. **Fig 8**. Note that the frequency and total doses are given from the booster dose onwards.

| Label | Booster | Frequency | Total doses |
| --- | --- | --- | --- |
|  | *standard = 3 weeks, early = 1 week* |  |  |
| 0 | standard | 3 weekly | 10 |
| s1 | standard | 1 week | 10 |
| s2 | standard | 3 weeks | 5 |
| s3 | standard | 14 weeks | 2 |
| s4 | standard | 1 week | 30 |
| s5 | standard | 3.5 days | 40 |
| e1 | early | 2 weeks | 10 |
| e2 | early | 1 week | 10 |
| e3 | early | 3 weeks | 5 |
| e4 | early | 1 week | 20 |
| e5 | early | 3.5 days | 40 |

### Case study: virtual cohort representative of human responses

To examine the robustness of our model and its predictions for a clinical setting, we generated a virtual cohort using the Phase II clinical trial of T-VEC [67]. As no in-human data of MV-BiTEs therapy exists, we used a study of an advanced oncolytic virus encoding an immunostimulatory agent as a surrogate, here a herpes simplex type 1-derived OV encoding GM-CSF, as a surrogate. Melanoma was the targeted tumour entity in both the pre-clinical MV-BiTEs study and the T-VEC trial. In this clinical trial, an intratumoural injection of *V_d_*_1_ = 10^6^ was administered initially, followed by fortnightly booster injections of *V_d_*_2_ = 10^8^ starting 3 weeks after the initial T-VEC injection. The percentage change from baseline (CFB) for 98 patients was then recorded (Fig 8A). We assumed the same dosage size for MV-BiTEs as used for T-VEC in the clinical trial and included the addition of BiTEs in the injection to match our *in vivo* experiments. We fixed the final time of the simulation as *T* = 23 weeks (*T* = 161 days) and measured the resulting tumour size *U* (*T*) = *U* (161).

We first examined whether heterogeneity in the initial tumour burden could explain the change from baseline (CFB) measurements in the clinical trial. To do this, we parameterised a gamma distribution using summary statistics reported by Dessinioti *et al.* [75] for the diameter of melanomas in 537 cases (see Methods and Supplementary Information for details). We then sampled 400 virtual patient initial tumour diameters (Fig 8B) and converted these to initial cell counts to feed into the model. The resulting CFB predicted from the model for heterogeneous initial tumour sizes was unable to capture the range of the clinical trial measurements (Fig 8C). This suggests that changes to parameters, representing patient-specific characteristics, are needed to capture clinical outcomes.

We next chose to follow Method (2), (Fig 3) to generate a cohort of 400 virtual patients (Method (2), Fig 3). We sampled 400 CFB to create a larger sample of human patients and ran a genetic algorithm to determine optimal parameter sets that matched the sampled CFB (S8 Fig). We fixed the initial tumour size to the median melanoma diameter and chose three parameters to optimise, *k, d_B_*, and *r* (Fig 8D), based on their sensitivity so far revealed in our analysis. The genetic algorithm returned a close match between individual virtual patient samples and the model (Fig8E-F). The distributions for the change from baseline of the real patient cohort and virtual patient cohort qualitatively match and passed the Kolmogorov-Smirnov test for similarity. We then examined the correlations between patient parameters and their resulting change from baseline (Fig 8D and S8 Fig). We found a particularly strong correlation between the tumour growth rate, *r*, and the T cell killing rate, *k*, with low growth rates and strong immune responses showing the largest negative CFB. The clearance of BiTEs, *d_B_*, also seems to positively correlate with CFB suggesting that a faster clearance of BiTEs results in a larger tumour size.

### Case study: impact of alternate treatment protocols on *in silico*

### human clinical trial

Lastly, we chose to examine whether an alternate, more effective, protocol exists. We generated 10 hypothetical treatment protocols that examined either the standard booster onset time (i.e. 3 weeks following the first *V_d_*_1_ dose) or an earlier booster onset time (i.e. 1 week following the first *V_d_*_1_ dose) and then varying intervals between *V_d_*_2_ doses and their multiplicity (Table 4 and Fig 8G). The total administered dose of MV-BiTEs, i.e. *V_T OT_* = 20 *× V_d_*_2_ was fixed across all protocols considered. Reduction in tumour size was largest under the standard booster onset (3 weeks after the first injection) with more injections given more regularly (s4 and s5, Fig 8H). In contrast, intervals between consecutive doses larger than the current standard fortnightly protocol, do not improve the average tumour burden (i.e. s1-s3, e1-e3). To examine the relationship between final tumour size and hypothetical survival curves for our virtual cohort, we have plotted the Kaplan-Meier survival curves under the 10 protocols for a survival threshold of 120% CFB (Fig 8I). These suggest that individuals predicted to have large tumour sizes early on under the standard protocol will benefit in the short-term from the early booster onset protocols with frequent smaller dosages (e4 and e5).

## Discussion

The potential therapeutic efficacy of BiTEs delivered by oncolytic viral vectors is an exciting area of immunotherapy. In this work, we develop a mathematical platform that can be used to simulate the efficacy of this novel therapeutic strategy. We show how this model recapitulates the dynamics of MV-BiTE *in vitro* and *in vivo*, as well as matches change from baseline (%) measurements in a Phase II clinical trial for T-VEC. Generating *in silico* clinical trials, we unravel the likely mechanistic drivers of the heterogeneity in responses to MV-BiTEs as well as propose potentially more effective protocols. This hypothesis-generating platform serves to better inform future experiments and, hopefully, the translation into clinical trials.

An overarching theme of this research has been to understand the impact of alterations to the MV-BiTEs administration protocol. Mathematical modelling is especially valuable to address this question, since it would be impossible (and unethical) to systemically test all conceivable perturbations in animal experiments. With our predictions, we can prioritise certain protocols for *in vivo* validation.

Despite good alignment of the model with our experimental data and despite delivering insightful simulations, this study has some limitations. Importantly, we do not expect MV-BiTEs therapy to yield the same results in a clinical trial as those in the Phase II T-VEC trial and instead use the results of this clinical trial as an example of heterogeneity that may be seen in an MV-BiTEs protocol. In the future, clinical data might be obtained from studies on BiTE-encoding OVs that more closely recapitulate the interactions described by the ODEs developed in this work. One such example is an ongoing clinical trial investigating an oncolytic adenovirus encoding several immunomodulatory transgenes including a FAP-targeting T cell engager [NCT04053283, [77]; NCT05043714, [78]]. Another caveat is the low number of individual mice the cohort simulation is based on, especially for the delayed tumour growth cohort 3 (n=1). Additional *in vivo* studies are expected to generate a more robust basis for our simulations, while dedicated measurements of, e.g., phagocytosis and antigen presentation can help to dissect additional aspects of immunological responses to OV-BiTEs therapy.

We found consistently throughout our simulations that lower-dosage injections given more frequently were predicted to reduce the tumour size most significantly. In both the *in silico* clinical trials based on the *in vivo* tumour growth and the Phase II clinical trial data, we saw that more frequent injections were optimal. Furthermore, we found that an initial dosage followed by a short delay in the following booster dosage (3 weeks) followed by frequent small doses was most effective. This suggests that once the treatment effect has time to establish after the first dosage, frequent small dosages are more effective than large bolus dosages given far apart. However, in experimental and clinical practice, this might pose a challenge due to tumour growth in the intervals between injections and, therefore, needs to be carefully evaluated in a well-designed animal experiment beforehand. Another caveat to this is that our model predicted individuals who would respond to MV-BiTE, would likely respond irrespective of the administration protocol. By contrast, those classified as experiencing tumour “growth” or having a large change from baseline (%) would benefit most from a change in protocol. This suggests that if it were possible to pre-determine which classification an individual patient would fall into, it would be possible to predict their potential success under treatment with MV-BiTEs.

In both *in silico* clinical trials generated, CD3^+^ T cell cytotoxicity and BiTE pharmacokinetic clearance were predicted to determine whether the tumour regressed or grew. This is in line with previous clinical findings that identified pre-existing T cell responses towards measles virus and tumour antigens to correlate with outcome of MV therapy [79]. *In vivo* experiments [13] as well as the mathematical model showed that encoding BiTEs in the MV vector is crucial for therapeutic effectiveness as compared to direct BiTE injections. Nevertheless, simulations indicated that a high BiTE clearance rate limited therapeutic success. This points towards temporary BiTE effects despite vector-mediated expression. Differences in BiTE pharmacokinetics could be due to individual immune responses (e.g., via neutralising antibodies or phagocytosis), variations in tumour composition (e.g., areas of hyperproliferation, hypoxia, or necrosis), or restrictions in the tumour microenvironment such as dense stroma. Predicting potentially crucial parameters may prove helpful in the search for biomarkers of response. If candidate parameters could be measured prior to treatment onset, e.g., levels of neutralising antibodies, they can be correlated with the predicted treatment outcome for validation. If this holds true, stratification into biomarker-adapted treatment arms could improve overall outcome.

In addition, our findings suggest that targeting BiTE pharmacokinetics could be one next avenue forward for experimental design, e.g., testing other, potentially more stable protein formats that have been proposed, such as DARTs, diabodies, or IgG-like structures [80, 81]. In addition, alternative vectors have been considered for delivery of BiTEs, such as other OVs [12, 15], (CAR) T cells [82, 83], or nanocarriers that deliver *in vitro*-transcribed mRNA encoding BiTEs to host myeloid cells [84]. Comparing the pharmacokinetics of these vectors could lead to improved efficacy at the clinical level. Importantly, our findings in patient-derived xenograft models with tumour cells of human origin showed extended BiTE expression compared to the murine tumour model, indicating that limited MV-BiTE effectiveness can, at least in part, be attributed to restricted viral replication in murine cells [13], giving hope for better outcomes in human patients.

There are certain limitations in our model based on some simplifying assumptions. For example, we assumed the growth of the control cell population in the cell viability measurements was negligible (see Supplementary Material). The growth of the cells *in vitro* would have likely been highly non-linear with periods of growth, saturation and then decline. Future work will investigate this limitation further by generating data for the cell viability measurements that may be used to better calibrate the model. We are confident, however, that the model still provides a sufficient baseline for approximating the impact of viral infection and death of tumour cells as a large number of parameters were recalibrated to the *in vivo* data.

Furthermore, a wide range of biological processes is involved in anti-tumour T cell effects, including activation via the endogenous T cell receptor and/or bispecific T cell engagers, replication of individual T cell clones, stimulation and inhibition by cytokines and other mechanisms, and T cell exhaustion. While our model captures rates of T cell accumulation, decay, and baseline as well as BiTE-induced tumor cell killing, it does not explicitly account for T cell exhaustion, which is a common mechanism found in cancer and, more specifically, in both OV and BiTE therapies. Various concepts such as OV-encoded immune checkpoint inhibitors or BiTE treatment-free intervals have been devised to address this [85–88]. In a future version of the model, we aim to implement T cell exhaustion on the basis of time-resolved functional T cell characterisation to better reflect cases of treatment failure under continuous BiTE release by OV-infected cells. This can subsequently be used to predict optimal combination or dosing strategies for long-term anti-tumour efficacy with specific focus on functional exhaustion of T cells.

In addition, MV-BiTEs can form syncytia which is omitted from our model. With the addition of data quantifying the presence of syncytia, we could extend the model to capture the effect of this on viral spread as is done in other mathematical models [89, 90].

Taken together, our model serves as a platform for future research in OV-BiTEs to be conducted and hypotheses to be tested. In particular, testing of more stable bispecific protein formats and investigation of selected treatment protocols will be conducted as experimental validation. Such validation is crucial for any modelling prediction.

Thereby, we aim at refining the model using novel datasets and, ultimately, at further developing complex combination immunovirotherapies towards clinical translation.

## Supporting information

**S1 Text** Details on the parameter fitting, parameter sensitivity and supporting analysis of the virtual cohort parameters.

**S1 Fig. Model simulation of the fit to cell viability and viral progeny measurements from B16 cells using Eq. S1.** The fits (solid lines) for (A) cell viability and (B) viral progeny, are shown against the individual experiment measurements (circle). Note in (B) the same fit is presented on a linear (left) and log (right) plot.

**S2 Fig. Fitting data from LDH release assay to obtain an estimate for BiTE-mediated tumour cell killing by T cells, i.e.** *ɛ, k* **and** *γ*. The mean and standard deviation for specific lysis % for varying BiTE concentrations is plotted in black. The model approximation using Eq. S2 is plotted in blue.

S3 Fig. Local pair-wise parameter perturbation by *p*^ **giving the relative total tumour cells on day 25 (Eq. S3) is represented by the colour bar.** Direction of perturbation is increasing moving down vertically and across horizontally for each parameter value.

**S4 Fig. Four individual parameter sets and their resulting model trajectories against m4’s data. These four-exemplar parameter sets, and their model trajectories are a subset of those plotted on Fig 5**. The black line and solid circles are the tumour volume measurements, and the orange curve is the model’s predicted solution.

**S5 Fig. Sampled parameters that give model trajectories matching the data for individual mice m1-m8 (Method (1), Fig 3).** Sampled 250,000,000 parameter sets and calculated the residual to each of the 8 mice’s tumour volume, i.e. m1 through to m8. The 400 parameter sets giving the lowest residual have been plotted as histograms for *k, ɛ, d_B_, d_T_, s, η* and *β* with their trajectories against the data of individual mice, mean values and standard deviations in Fig 5.

**S6 Fig. Pairwise scatter plots for significant parameter correlations across the 8 mice, m1-m8.** The *x, y* labels denote the parameters of interest, and the titles denote the mouse. Points correspond to the specific parameter values for that mouse. The colour of points corresponds to the mouse.

**S7 Fig. Impact of the number of injections and days between injections of MV-BiTEs for the three cohorts: 1) recovery, growth and delayed-growth, corresponding to Fig 7**. The corresponding mean and standard deviation for the three cohorts (c1, c2, and c3) at (A-B) 48 days and (C-D) 7 days after the last injection.

**S8 Fig. Additional information about the virtual cohort expanded from human clinical trial data.** (A) Change from baseline (%) for the Phase II T-VEC clinical trial digitized using WebPlotDigitiser. (B) Probability density function (PDF) generated for the clinical trial data and compared to the virtual clinical trial (VCT) samples. (C) The resulting change from baseline for the model’s optimised parameters using a genetic algorithm. (D) Correlation matrix for the three patient-specific parameters sampled to match the final tumour size of the virtual patients *U* (*T*).

## Data Availability Statement

The datasets generated for this study can be found in the https://github.com/AdrianneJennerQUT/insilico-clinical-trial-MV-BiTEs

