## Supplementary Information for "In silico clinical trials of BiTE expression by oncolytic viruses reveal the impact of patient heterogeneity on dosage protocol"

#### 1 Details of the parameter fitting for the model

Parameters in **Eqs. 1-5** were fit to *in vitro* and *in vivo* measurements from Speck *et al.* [1] and measurements generated for this study. There were five datasets used to fit parameters in the model: viral progeny, cell viability, BiTE fold change, specific lysis (from [1]) and *in vivo* tumour volume. Details of the experiments can be found in the main text Methods section. The parameters fit to each experiment are summarized in **Table 2** with model variables in **Table 1** in the main text.

##### 1.1 Fitting algorithm details

Parameters were fit using a nonlinear least square fitting algorithm. Briefly, consider a vector of $n$ parameters $\mathbf{p} = [p_1, p_2, \dots p_n]$ to fit to a vector of $m$ data points $\mathbf{y} = [y_1, y_2, \dots y_m]$ corresponding to a set of time points $\mathbf{t_d} = [t_1, t_2, \dots, t_m]$ where $m \gg n$. The model $M(\mathbf{p}, t)$ (**Eqs. 1-5**) is a function of the parameters and time. Consider the vector of outputs of our model that correspond to the vector of data points as $M(\mathbf{p}, \mathbf{t_d})$, then our objective is to minimize the following:

$$\min_{\mathbf{p}} ||\mathbf{y} - M(\mathbf{p}, \mathbf{t_d})||^2$$

i.e. to minimize the sum of squares error between our model approximation and the data. In some cases, to improve the estimation of the parameters, we fit multiple experiments simultaneously [2,3]. In the case of the simultaneously fit data sets, the vector of data points $\mathbf{y}$ then consists of data from two or more experiments and the model used would depend on the experiment.

In addition to this, we used a multi-start algorithm to sample multiple initial guesses for our parameter set. When run, the solver attempts to find multiple local solutions to a problem by starting from various points, and then from this obtains a global optimal solution. This means the nonlinear-least squares fitting algorithm was run from multiple points within a pre-set parameter interval, which we set to be quite large, based on the parameter being fit. All multi-start algorithm runs resulted in a global optimum for our model fitting.

##### 1.2 Fitting parameters to the *in vitro* data set

###### 1.2.1 Viral progeny and cell viability

The viral progeny and cell viability measurements from [1] were used to fit the viral kinetic parameters $r, \beta, \alpha_V, \eta$ and $d_V$. As there was no immune system present in these *in vitro* experiments, and the impact of BiTEs was negligible in the absence of T cells, this reduced **Eqs. 1-5** to:

$$\frac{dU}{dt} = rU - \frac{\beta UV}{U+\eta},$$
$$\frac{dV}{dt} = \frac{\beta UV}{U+\eta} - d_V V, \qquad \text{S1}$$

where the total viruses were $V^* = \alpha_V V$. To fit the cell viability, we first assumed that the cell count for the cells in the control experiment would be constant, i.e. $U(t) = U(0)$. In reality, cells in the control experiment would likely grow initially, reach a plateau and then decrease. However, to conserve the simplicity of our model in **Eq. S1,** we chose to assume that this growth was negligible and could be assumed to be zero, i.e. $dU/dt = 0$, as the main kinetic of interest was the impact of the virus. Furthermore, in the absence of quantitative data, i.e. absolute cell numbers for the control experiment, it is not mathematically possible to obtain with certainty an estimate for the growth function of the control cells. This gave the following equation for calculating cell viability:

$$\text{cell viability} = \frac{U(t)}{U(0)} \times 100.$$

We fit all remaining parameters in the model to the data. We also expect $r$ to be small given the constraints of proliferating in the dish when close to confluence. The initial conditions were fixed as $U(0) = 10^5\ cells$ and $V(0) = 10^3$ cells. The fitted parameter values are in **Table 2** in the main text. The resulting fit can be found in **Figure S1**~~**Figure S1**~~.

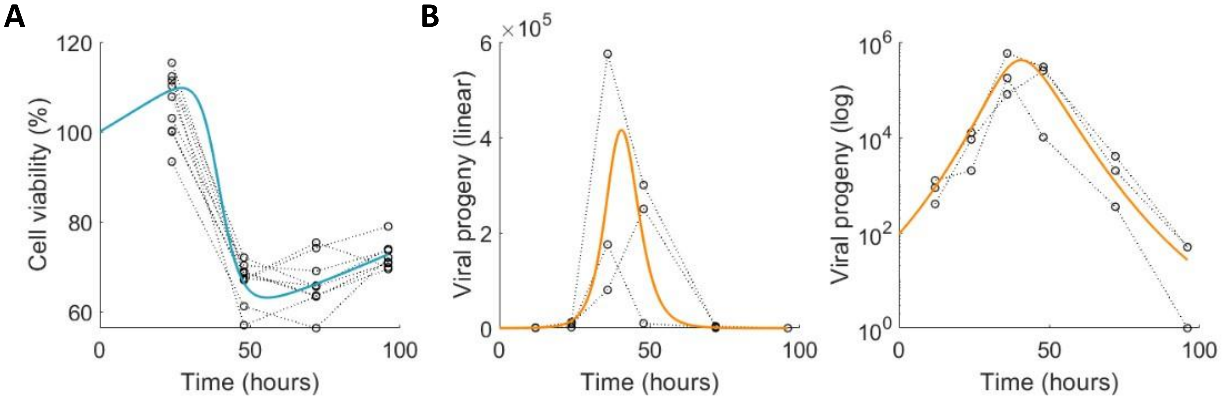


**Figure S1 Model simulation of the fit to cell viability and viral progeny measurements from B16 cells using Eq. S1**. The fits (solid lines) for **(A)** cell viability and **(B)** viral progeny, are shown against the individual experiment measurements (circle). Note in **(B)** the same fit is presented on a linear (left) and log (right) plot.

##### 1.2.2 BiTE %specific lysis

To estimate the impact of BiTEs on T cell mediated tumour cell killing, we used the % specific lysis values from LDH assays from [1] to estimate $\epsilon$, $k$ and $\gamma$. As specific lysis is calculated using the number of cells that have died $D(t)$ and since there was no virus present in this experiment, we could reduce the system to

$$\frac{dU}{dt} = rU - \frac{kTU}{N}\left(\epsilon + \frac{B}{B+\gamma}\right),$$
$$\frac{dD}{dt} = \frac{kTU}{N}\left(\epsilon + \frac{B}{B+\gamma}\right), \qquad \text{S2}$$
$$\frac{dB}{dt} = 0,$$
$$\frac{dT}{dt} = 0.$$

Fitting the data, we fit to

$$\frac{D(48)}{D(48) + U(48)}.$$

Note, that since we did not have any measurements for the decay of BiTEs or immune cells over the 48-hour period, we assumed that there was no decay in this *in vitro* setting, i.e. $d_B = 0$. In the *in vivo* setting, we would expect the decay rate of BiTEs and immune cells to be non-negligible. We also assumed that in the time frame of the experiment, the death rate of dead cells would be negligible, hence $d_D = 0$.

We fixed the parameters in the model that had been fitted in **Figure S1** and set the initial conditions to be $U(0) = 10^5, D(0) = 0,$ and $T(0) = 2.5 \times 10^5$. The initial concentration of BiTEs was then given in the experiment, i.e.$B(0) = [1, 0.1, 0.01, 0.001, 0.0001, 0]\ \mu g/mL$. The fit to the data is given in **Figure S2**.

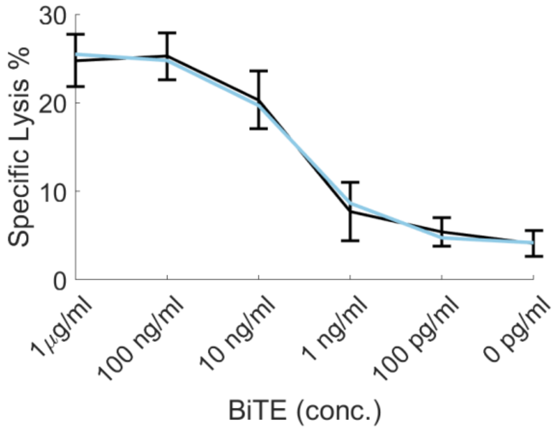


**Figure S2. Fitting data from LDH release assay to obtain an estimate for BiTE-mediated tumour cell killing by T cells, i.e. $\epsilon$, $k$ and $\gamma$.** The mean and standard deviation for specific lysis % for varying BiTE concentrations is plotted in black. The model approximation using **Eq. S2** is plotted in blue.

##### 1.3 Complete *in vivo* dataset

Tumour volume was measured *in vivo* after (A) control (mock), (B) purified BiTEs only, (C) MV only, and (D) MV-BiTEs, see **Figure 4 i**n the main text. As mice were sacrificed during the experiment when their tumour reached a given threshold, the cohort number changes over time. To avoid fitting to unrealistic mean population data, we only use data while more than 5 mice (of the $n = 10$ per group) were still alive.

##### 1.3.1 *In vivo* tumour growth fitting (control)

Until now, we had been fitting *in vitro* experiments and assuming the underlying tumour growth under treatment in the short time frame (48 – 96 h) was exponential. For the *in vivo* mouse model and human patient simulations, we needed a more accurate estimate of tumour growth. Assuming a Gompertzian tumour growth model, we fit the growth rate $r$ and carrying capacity $L$ for a tumour to B16-CD20-CD46 *in vivo* control tumour growth data: $\frac{dU}{dt} = rU\log\left(\frac{L}{K}\right)$ (**Figure 4B**). To convert the number of cells to a volume and fit to the data we assumed:

$$volume = \frac{U(t)}{10^6}$$

i.e. $10^6$ $cells$ per $mm^3$, as has been done by other authors previously [5,6]. Note, while we know there are immune cells present, due to the absence of BiTEs and MV we assume their effect on tumour growth is negligible and hence $T(t) = 0$.

##### 1.3.2 *In vivo* tumour growth (BiTEs, MV and MV+BiTEs)

To fit the remaining model parameters, we simultaneously fit all remaining *in vivo* tumour growth measurements (BiTEs, MV, and MV-BiTEs). In other words, we minimised the distance between the model and the three data sets (see Section 1.1 above) assuming all parameters were fixed across the experiments. Variations on the full model were used based on the presence/absence of BiTEs or virus:

| **BiTE** | **MV** |
|---|---|
| $\frac{dU}{dt} = rU\log\left(\frac{K}{U}\right) - \frac{kUT}{N}\left(\epsilon + \frac{B}{B+\gamma}\right),$ <br> $\frac{dD}{dt} = \frac{kUT}{N}\left(\epsilon + \frac{B}{B+\gamma}\right) - d_D D,$ <br> $\frac{dB}{dt} = -d_B B,$ <br> $\frac{dT}{dt} = sD - d_T T$ | $\frac{dU}{dt} = rU\log\left(\frac{K}{U}\right) - \frac{\beta UV}{U+\eta} - \frac{kUT}{N}(\epsilon),$ <br> $\frac{dD}{dt} = \frac{\beta UV}{U+\eta} + \frac{kUT}{N}(\epsilon) - d_D D,$ <br> $\frac{dV}{dt} = \frac{\beta UV}{U+\eta} - d_V V,$ <br> $\frac{dT}{dt} = sD - d_T T$ |

| **MV-BiTE** |
|---|
| $\frac{dU}{dt} = rU\log\left(\frac{K}{U}\right) - \frac{\beta UV}{U+\eta} - \frac{kUT}{N}\left(\epsilon + \frac{B}{B+\gamma}\right),$ <br> $\frac{dD}{dt} = \frac{kUT}{N}\left(\epsilon + \frac{B}{B+\gamma}\right) - d_D D,$ <br> $\frac{dV}{dt} = \frac{\beta UV}{U+\eta} - d_V V,$ <br> $\frac{dB}{dt} = -d_B B,$ <br> $\frac{dT}{dt} = sD - d_T T$ |

The following parameters were fit: $s, d_D, \beta, d_B, \epsilon, \alpha_B$ and $k$, with resulting fits in **Figure 4C-E**. The number of tumour cells initially was $U(0) = 10^6$ and the number of immune cells present was fixed to $T(0) = 2.5 \times 10^5$ cells. In the BiTE only group, a BiTE dose of $2.17\mu g$ (in $100\mu L$ suspension) was injected on day one, yielding a concentration of 21.7 g/ml . In the MV only group, a MV dose of $10^6$ infectious units was injected every day for 5 days. In the MV-BiTE group, MV encoding BiTE was injected along with the BiTE dosage from the BiTE experiment (21.7 g/ml). This gave the initial conditions: $B(0) = 21.7\mu g/ml$ and $V(0) = 10^6$ [cell] infectious units.

#### 2 Local parameter sensitivity analysis

To investigate the sensitivity of the parameterised model, we conducted a local (**Figure S3**) and global (**Figure 4F**) parameter sensitivity analysis. We perturbed pairs of parameters from their fitted value by a factor of

$$\hat{p} = [0.7, 0.8, 0.9, 1, 1.1, 1.2, 1.3],$$

and measured the resulting relative tumour size on day 25 from a single injection of MV-BiTEs compared to baseline $U_B$:

$$\frac{U(25) - U_B(25)}{U_B(25)}. \qquad \text{S3}$$

We chose a final day of day 25 as this was close to the range of time for the average tumour volume measurements *in vivo*. Results of this are presented in **Figure S3.** The local sensitivity analysis showed that the viral infectivity, $\beta$, and the clearance of virus $d_V$ were the most sensitive to perturbations.

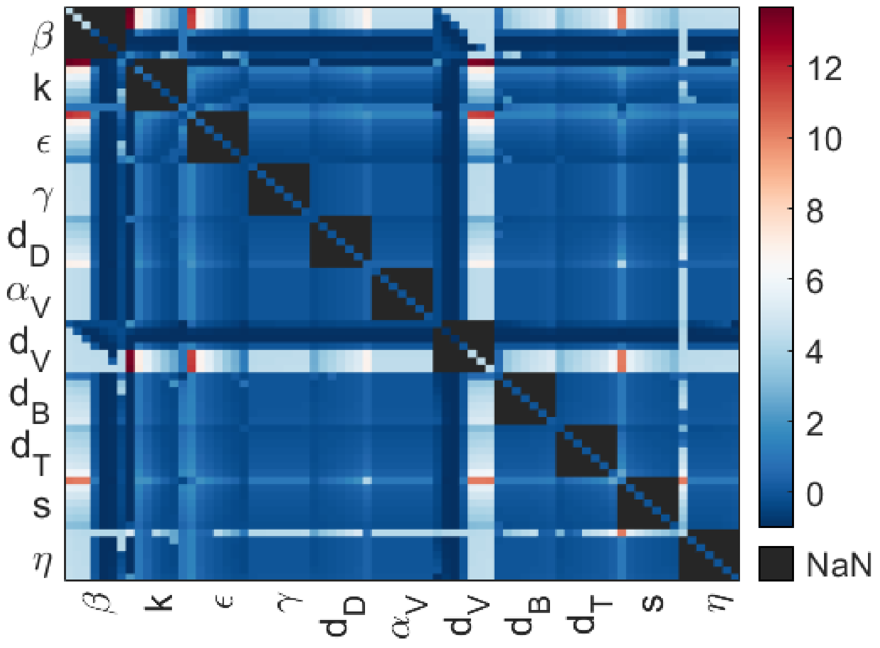


**Figure S3 Local pair-wise parameter perturbation by $\hat{p}$ giving the relative total tumour cells on day 25 (Eq. S3) is represented by the colour bar.** Direction of perturbation is increasing moving down vertically and across horizontally for each parameter value.

#### 3 Rescaling of parameters for clinical trial simulation

As both $L$ and $\eta$ have cells as unit and were estimated *in vivo*, these were rescaled to a human context for use with the human clinical trial data. We took the largest change from baseline in the clinical trial data set which was 974%. The initial tumour size in the clinical trial was assumed to be $U(0) = 1.15 \times 10^9$ cells. Assuming the largest increase is the carrying capacity

$$L = \frac{975}{100} \times U(0) + U(0) = 5.6 \times 10^9.$$

We then scaled $\eta$ by the increase from the initial number of tumour cells in the *in vitro* experiment (i.e. $10^6$) to the new initial condition for the human cohort, i.e. $U(0) = 1.15 \times 10^9$

$$\eta = 9.04 \times 10^5 \times \frac{U(0)}{10^6} = 4.7 \times 10^8$$

And similarly for $\alpha_B$ (recall the units in **Table 2**):

$$\alpha_B = 4.6 \times 10^4 \times \frac{10^6}{U(0)} = 87.85.$$

#### 4 Parameter values and correlations for virtual cohort matching individual trajectories

In a first attempt to capture the inherent heterogeneity in the tumour volumes of individual mice, we generated a virtual cohort using Method (1) (**Figure 3**) for each individual mouse m1-m8. An example of four individual parameter sets and their resulting trajectories against m4's data is given in **Figure S4.** The resulting histograms for the 400 parameter samples for each mouse are given in **Figure S5** with corresponding trajectories, mean values and standard deviations in **Figure 5**. Histograms summarising the parameter values obtained for each individual mouse for each model parameter are shown in **Figure S6**. Pair-wise parameter scatter plots in **Figure S7** are provided to show how some parameters correlate.

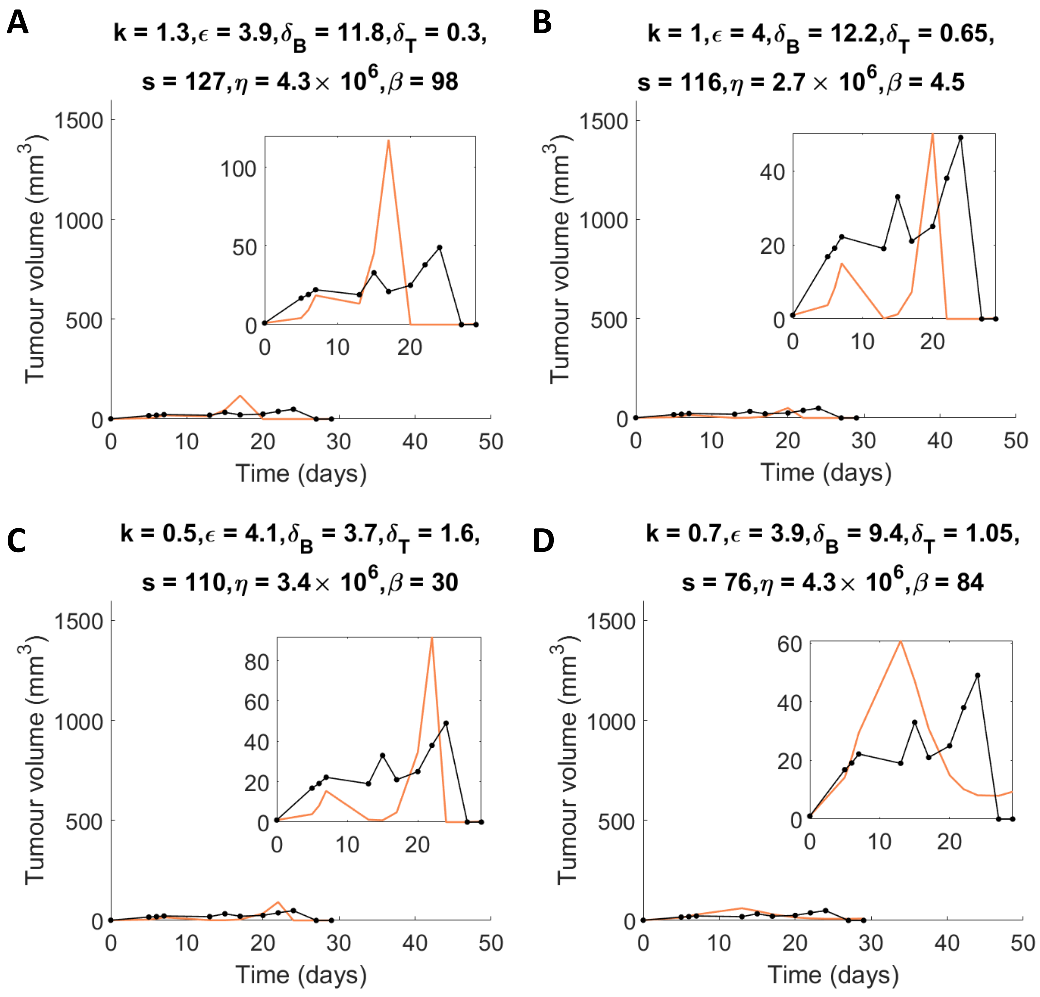


**Figure S4 Four individual parameter sets and their resulting model trajectories against m4's data.** These four-exemplar parameter sets, and their model trajectories are a subset of those plotted on **Figure 5.** The black line and solid circles are the tumour volume measurements, and the orange curve is the model's predicted solution.

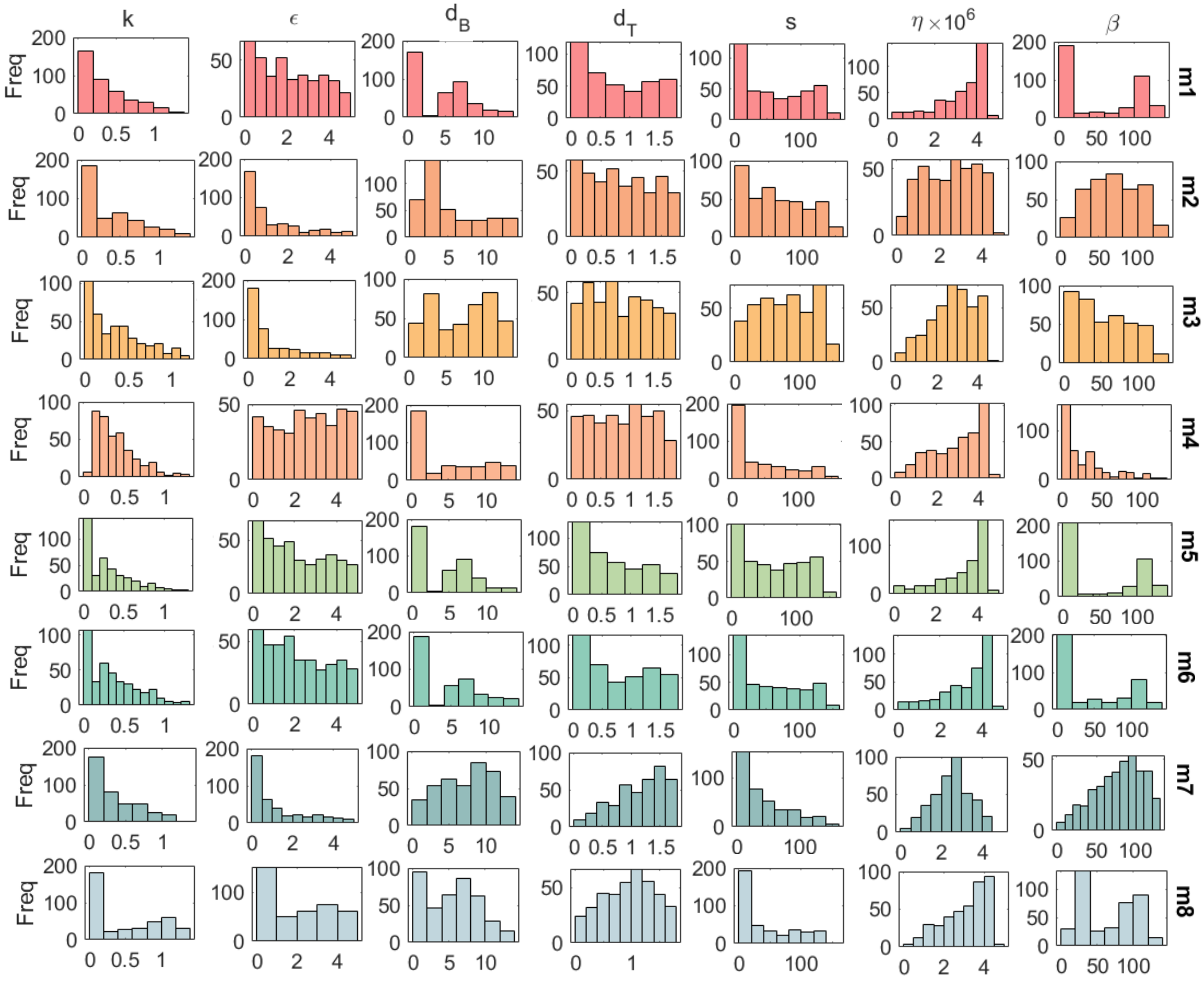


**Figure S5 Sampled parameters that give model trajectories matching the data for individual mice m1-m8 (Method (1), Figure 3).** Sampled 250,000,000 parameter sets and calculated the residual to each of the 8 mice's tumour volume, i.e. m1 through to m8. The 400 parameter sets giving the lowest residual have been plotted as histograms for $\boldsymbol{k}, \boldsymbol{\epsilon}, \boldsymbol{d_B}, \boldsymbol{d_T}, \boldsymbol{s}, \boldsymbol{\eta}$ and $\boldsymbol{\beta}$ with their trajectories against the data of individual mice, mean values and standard deviations in **Figure 5**.

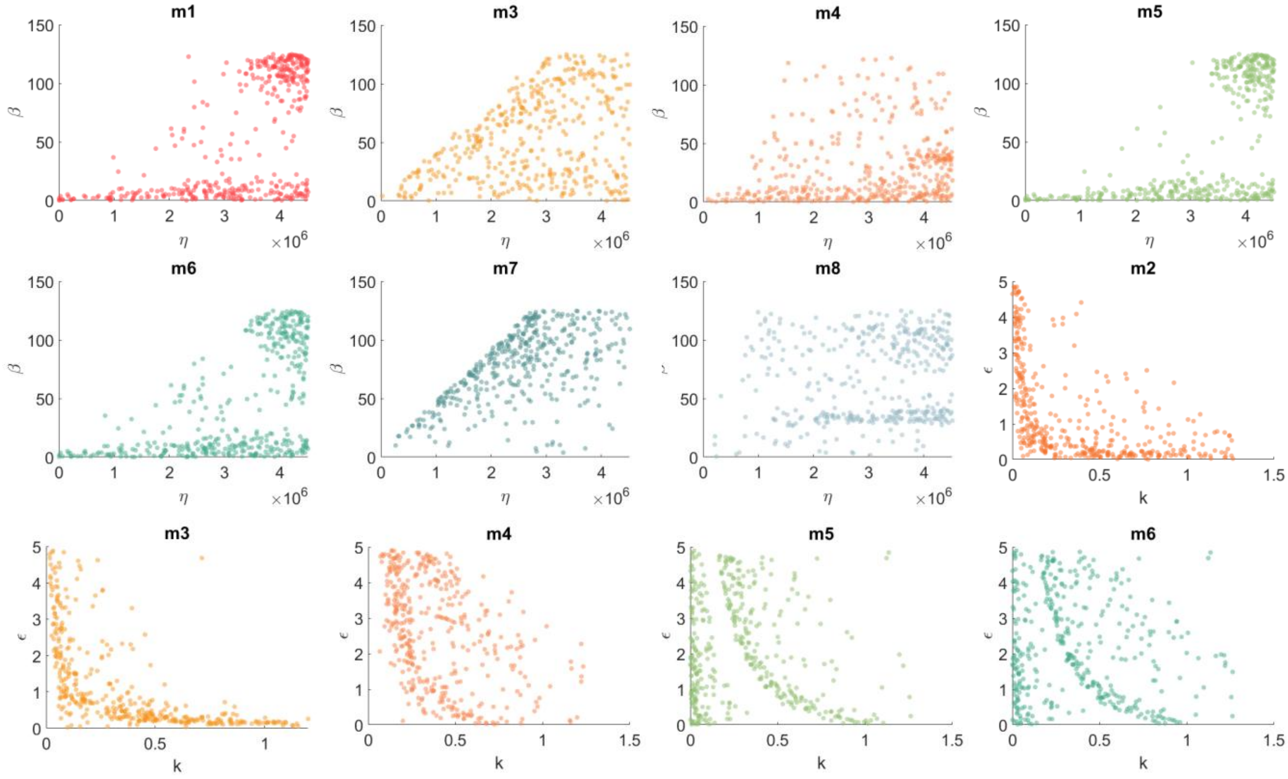


**Figure S6 Pairwise scatter plots for significant parameter correlations across the 8 mice, m1-m8.** The x,y labels denote the parameters of interest, and the titles denote the mouse. Points correspond to the specific parameter values for that mouse. The colour of points corresponds to the mouse.

#### 5 Virtual cohorts clustered by growth, delayed growth and recovery

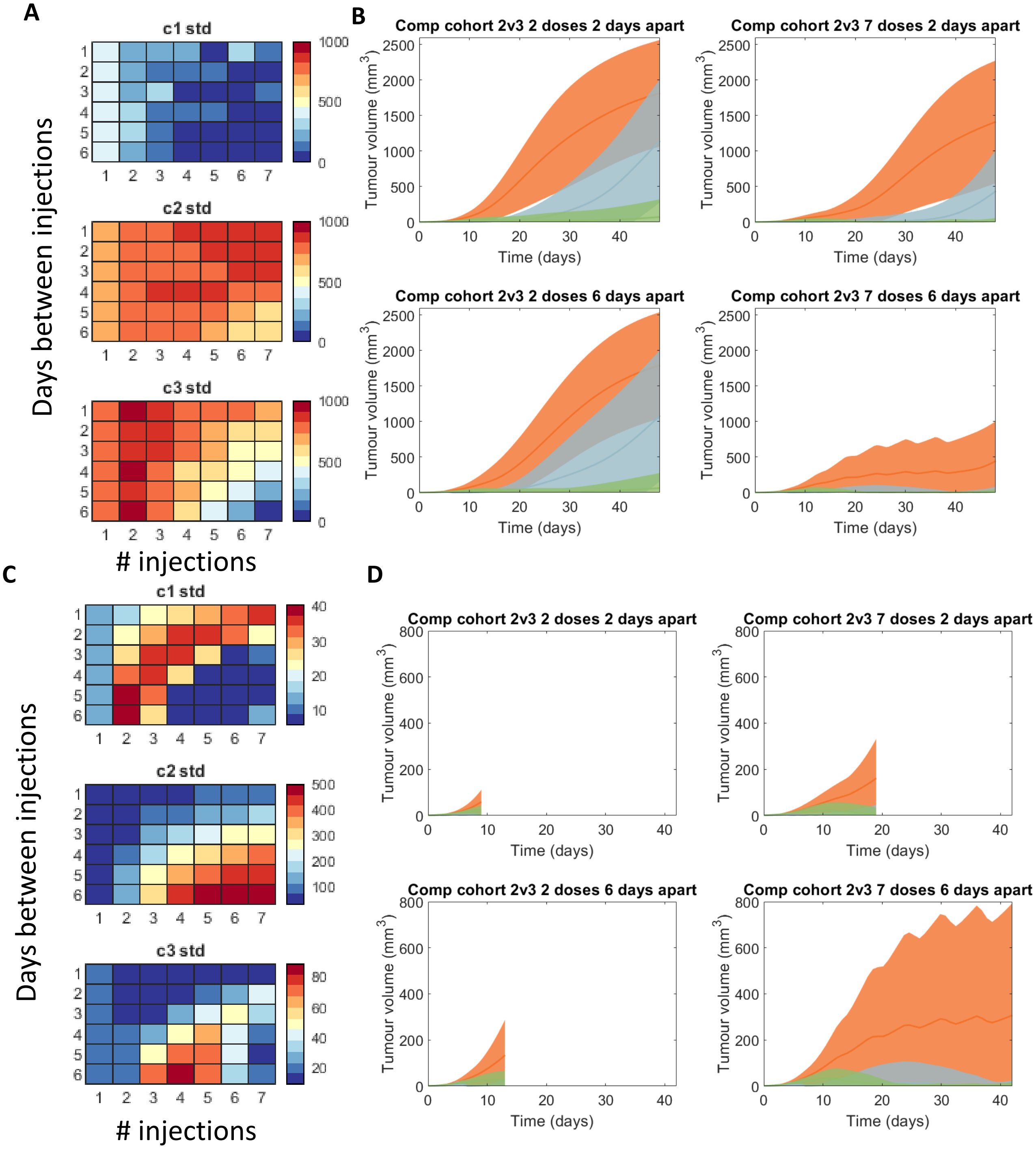


**Figure S7 Impact of the number of injections and days between injections of MV-BiTEs for the three cohorts: 1) recovery, growth and delayed-growth, corresponding to Figure 7**. The corresponding mean and standard deviation for the three cohorts (c1, c2, and c3) at **(A-B)** 48 days and **(C-D)** 7 days after the last injection.

### 6 Generating a virtual cohort based on T-VEC Phase II human clinical trial

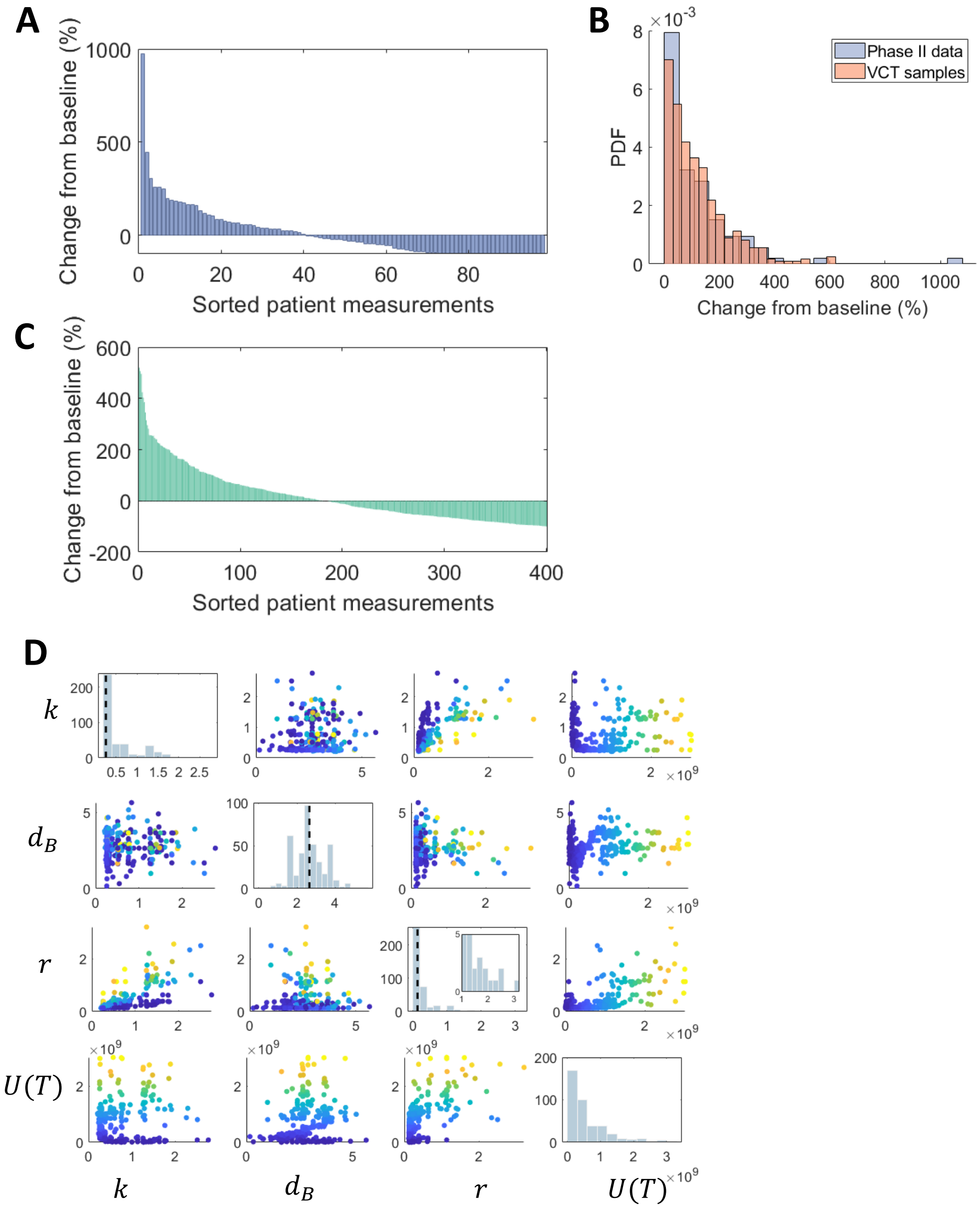


**Figure S8 Additional information about the virtual cohort expanded from human clinical trial data**. **(A)** Change from baseline (%) for the Phase II T-VEC clinical trial digitized using WebPlotDigitiser**. (B)** Probability density function (PDF) generated for the clinical trial data and compared to the virtual clinical trial (VCT) samples. **(C)** The resulting change from baseline for the model's optimised parameters using a genetic algorithm. **(D)** Correlation matrix for the three patient-specific parameters sampled to match the final tumour size of the virtual patients $\boldsymbol{U(T)}$.

#### 7 Defining a distribution for initial melanoma sizes to sample from for virtual patients

Dessinioti *et al.* reported from 537 cases, melanomas had a median diameter of 13mm, an interquartile range of 9mm to 20mm and 10.6% had a diameter of less than 6mm (see references in the main text). Given that melanomas will have strictly positive diameter, we assumed samples for our melanoma diameters should come from a gamma distribution, i.e.

$$f(x) = \frac{1}{\Gamma(k)\theta^k} x^{k-1} e^{-\frac{x}{\theta}}$$

where $k$ is the shape parameter and $\theta$ is the scale. Recall, that the cumulative density function (CDF) CDF is defined as

$$F(x) = P(X \leq x)$$

Where $F(x)$ represents the probability of $X$ being less than of equal to $x$. For example, for $10.6\%$ of measurements to be less than 6mm, then $F(6) = 0.106$. To estimate $\alpha$ and $\theta$, we minimise the error between the statistical measurements (median, 10.6%, and interquartile ranges) and the gamma function CDF using MATLAB's *gaminv*. For example, for 10.6% we calculate the error as $6 - F^{-1}(10.6)$, where $F^{-1}$ is the inverse CDF. From this we get the shape and scale parameters, $k = 3.09, \theta = 4.91$ and the corresponding distribution of samples in Fig 8B (main text).
